# Circulating plasma microRNAs miR-150 and miR-375 levels are associated with age-related endotypes of newly diagnosed Type 1 Diabetes

**DOI:** 10.64898/2026.02.18.26346540

**Authors:** Giuseppina Emanuela Grieco, Erika Pedace, Giada Licata, Tomi Suomi, Inna Starskaia, Laura L. Elo, Tim Tree, Riitta Lahesmaa, Pia Leete, Sarah J. Richardson, Noel G. Morgan, Francesco Dotta, Guido Sebastiani

## Abstract

Age-defined type 1 diabetes (T1D) endotypes, T1DE1 and T1DE2, are characterized by reproducible differences in pancreatic immunopathology and clinical course. In particular, these endotypes differ in the extent and composition of lymphocytic insulitis and in the extent of loss of insulin-producing β cell mass, at diagnosis. However, blood-based biomarkers that may distinguish these endotypes and inform the underlying immune–islet biology axis at diagnosis remain limited. Here, we characterized the clinical features and profiled circulating microRNAs (miRNAs) in plasma from two independent INNODIA cohorts of individuals with newly diagnosed stage 3 T1D (discovery, n=115; replication, n=147), stratified into age-defined endotypes (T1DE1, <7 years; T1DE2, ≥13 years; and intermediate T1DInt, 7–12 years). Differential-expression and age-adjusted models were coupled to orthogonal ddPCR validation. Putative miRNAs cellular sources were inferred using reference miRNA expression atlases. Biological context was explored via correlations of miRNAs with whole-blood transcriptomics.

Clinically, T1DE1 was associated with lower β-cell function and higher first-year C-peptide decline, alongside distinct islet autoantibody patterns, consistent with an immunologically aggressive endotype. Small RNA-seq analysis and ddPCR validation identified a reproducible signature in which miR-150-5p, a B-and T-lymphocyte related miRNA, and miR-375-3p, a β cell enriched molecule, were consistently increased in T1DE1 compared with T1DE2 across both cohorts. MiR-150-5p retained robust association with T1DE1 even after age adjustment, and neither miRNA was associated with age in non-T1D pediatric datasets, supporting T1D endotype specificity. The increased circulating miR-150-5p signal was not explained by differences in peripheral blood B-or T-cell frequencies in high-parameter flow-cytometry subsets, and its levels correlated inversely with whole-blood expression of the immune-associated miR-150-5p target genes MPPE1 and RABGAP1L.

Finally, applying a rule-based combined classifier (miR-150-5p and miR-375-3p “high”) achieved re-stratification of T1D individuals, including those in the intermediate age group, into two miRNA-defined groups with distinct β cell functional trajectories.

Collectively, these data suggest circulating miR-150-5p and miR-375-3p as non-invasive biomarkers linked to endotype-associated biology at T1D diagnosis, with potential utility for endotype-centered stratification and trial enrichment.

## INTRODUCTION

Type 1 diabetes mellitus (T1D) is a disease in which autoreactive immune responses progressively destroy insulin-producing pancreatic β cells, resulting in lifelong insulin dependence and chronic hyperglycaemia. While T1D often presents in childhood or adolescence, its onset can occur across the entire lifespan and the disease course varies markedly between individuals (1)(2). Accumulating evidence supports clinically meaningful heterogeneity in T1D pathogenesis, progression, and pancreatic pathology (3). Notably, age at diagnosis is associated with disease progression and particular immunometabolic features, suggesting the existence of distinct underlying pathogenic programs (4). Indeed, younger age at onset is correlated with an elevated genetic risk score, higher rate of progression, distinct metabolic and immunological patterns, and specific pancreatic histological features which, collectively, imply the existence of disease endotypes (5, 6). Consistent with this concept, pancreatic studies have identified two immunopathological endotypes, type 1 diabetes endotype 1 (T1DE1) and endotype 2 (T1DE2), that correlate with age at onset and differ in immune cell insulitis composition, inflammatory signalling, and β cell loss (7–11).

T1DE1 is typically associated with early onset of disease, often in childhood (age below 7y), and is characterized by rapid and extensive β cell destruction, leading to early and complete loss of insulin secretory capacity. In the pancreas, T1DE1 is characterised by lower β cell content, a high-grade lymphocytic infiltration of pancreatic islets, with a high proportion of CD20+ B cells and CD8+ T cells, alongside an interferon-stimulated gene (ISG) signature, indicative of an ongoing type I interferon response. This endotype reflects a more aggressive autoimmune process and has been linked to HLA-DR4 genetic background and the presence of multiple autoantibodies at diagnosis. In contrast, T1DE2, more frequent in individuals diagnosed at older ages (typically greater than 12y), exhibits a slower rate of β cell decline and preserved C-peptide levels for a longer duration. In the pancreas, T1DE2 is associated with a milder insulitis, often dominated by CD8+ T cells but with fewer infiltrating B cells and minimal interferon activity (7–9).

Despite these advances, scalable non-invasive biomarkers that capture endotype-associated biology at diagnosis remain limited. Importantly, although age at onset effectively stratifies patients into distinct endotypes, it does not capture the underlying tissue immunopathology and thus cannot serve as a biomarker of the immune landscape (12, 13). Therefore, identification of additional biomarkers that further distinguish the two endotypes and which help clarify the pathological mechanisms underlying their different characteristics and phenotypes is strongly needed. MicroRNAs (miRNAs), small non-coding RNAs that negatively regulate gene expression, have been addressed as useful blood circulating biomarkers in multiple diseases including T1D (14, 15). MiRNAs play crucial roles in immune regulation, influencing the differentiation, activation, and function of adaptive and innate immune cells (16), as well as β cell survival and function (17). Previously, we have shown that miRNAs can be secreted from CD4+ T lymphocytes and transferred to β cells during T1D (18); notably, miRNAs can also be secreted by pancreatic β cells under basal and/or stress/inflammatory conditions (19–22). Further, miRNA detected in whole blood very early on was associated with progression to beta-cell autoimmunity in young children (23).

Importantly, secreted miRNAs, detectable and measurable in blood plasma through a standardised protocol (24), can be helpful in distinguishing distinct clinical trajectories of different subgroups of T1D individuals (25, 26), or for predicting the response to immunomodulatory (27) or metabolic (28) treatments in diabetes.

Given the need for biomarkers that would help in refining the biological mechanisms underlying the two T1D endotypes (T1DE1 and T1DE2), we tested whether plasma miRNA profiles discriminate T1DE1 from T1DE2 and provide a blood-based readout of endotype-specific disease programs. We profiled circulating miRNAs in plasma from 262 individuals with newly diagnosed stage 3 T1D across two independent INNODIA cohorts to discover and validate endotype-associated miRNA signatures. We identified two miRNAs, miR-150-5p and miR-375-3p, that are increased in plasma of T1DE1 individuals compared with T1DE2. Particularly, these two miRNAs are preferentially expressed in B-and T-lymphocytes (miR-150) or in β cells (miR-375), with pivotal functional intracellular roles. Notably, these miRNAs were not associated with age in control cohorts and can be both used to re-stratify individuals with T1D across a population (including T1DEInt) into two distinct groups (miRC1, miRC2) independently of age at onset but with clinically distinct features resembling T1DE1 and T1DE2. Overall, these results suggest that these circulating miRNAs may capture the ongoing endotype biology in T1D.

## Results

### Age-defined endotypes used for study design in INNODIA T1D cohorts

In our previous study (25), using small RNA-sequencing, we profiled circulating miRNAs in plasma samples obtained from two independent INNODIA cohorts (first and second INNODIA cohort) of T1D individuals, enrolled within 6 weeks from diagnosis of stage 3 T1D in the INNODIA Natural History Study (29)(30).

In the present study, we re-analyzed these small RNA-seq datasets [GSE265980 (First cohort), GSE265981 (Second cohort)] to identify circulating miRNAs differentially expressed between endotype T1DE1 (age at onset < 7 years) and endotype 2 (T1DE2; age at onset ≥ 13 years). In both cohorts, stage 3 T1D participants were re-stratified into three groups, based on the age at onset: T1DE1 (<7y), T1DE2 (≥13y), and an intermediate group (potentially comprising individuals of either endotype: T1DE-Int (7-12y) (**Figure 1**).

**Figure 1.**
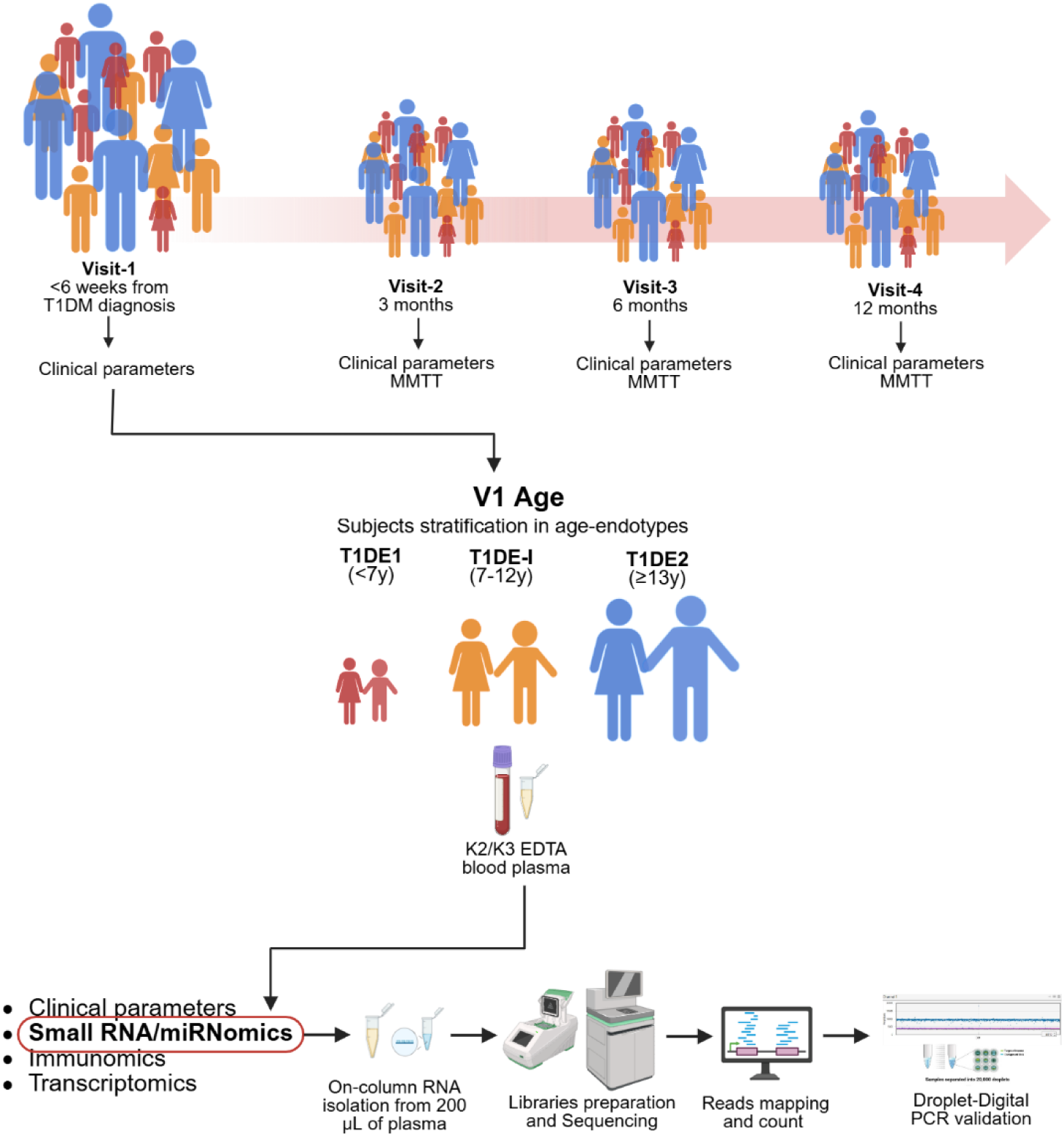
Study design, longitudinal follow-up, endotype stratification, and small RNA profiling workflow. Participants with stage 3 type 1 diabetes were followed longitudinally across four study visits from diagnosis: Visit 1 (V1, baseline; <6 weeks from diagnosis), Visit 2 (V2; 3 months), Visit 3 (V3; 6 months), and Visit 4 (V4; 12 months). Clinical parameters were collected at all visits, and mixed-meal tolerance tests (MMTT) were performed at V2, V3 and V4. At baseline (V1), participants were stratified into age-defined endotypes: T1DE1 (<7 years), T1DE-Int (7–12 years), and T1DE2 (≥13 years). Peripheral blood was collected in K_2_ or K_3_-EDTA tubes and processed to obtain plasma for downstream molecular profiling. The figure highlights the multi-layer phenotyping strategy, including clinical measurements and integrated-omics (small RNA/miRNomics, immunomics, and transcriptomics), with the primary molecular workflow shown for circulating small RNAs: on-column RNA extraction from 200 µL plasma, small RNA library preparation and sequencing, bioinformatic read mapping and quantification, and orthogonal validation of selected miRNA targets by droplet digital PCR (ddPCR).

The cohorts comprised n = 115 (58 female and 57 male, age 12.4 ± 7.7 years; mean T1D duration 4.5 ± 1.5 weeks) and n = 147 participants (55 female and 92 male; age 11.9 ± 7.9 years; mean T1D duration 3.9 ± 1.7 weeks) (**Table 1S**), respectively, each followed for 12 months with four scheduled visits (V1, baseline; V2, 3 months; V3, 6 months; V4, 12 months), at which key metabolic and immunological outcomes were assessed. Clinical outcomes at baseline and follow-up were compared across age endotypes. Differential-expression analyses of the miRNA-seq data were performed between T1DE1 and T1DE2. To minimize confounding by age per se, the following analyses were performed: (i) miRNAs differential expression analysis was repeated using age as a covariate in the model; (ii) candidate miRNAs were re-tested in multiple independent cohorts of controls without diabetes and individuals with other inflammatory/autoimmune diseases (i.e. asthma / celiac disease) but not type 1 diabetes, appropriately re-stratified by age (< 7 y, 7–12 y, ≥ 13 y). Additional-omics datasets (immunomics-transcriptomics) from the first and second INNODIA cohorts were further analyzed to explore potential associations between differentially expressed miRNAs across endotypes and other related immunological and molecular factors contributing to the biology of the two endotypes (**Figure 1**). Finally, the specific set of identified miRNAs was used to unbiasedly re-stratify the full population of T1D individuals.

**Table 1.** Demographics and clinical characteristics at Visit 1 (V1 - baseline) of Stage 3 T1D individuals of the INNODIA first cohort (n=115) subdivided into endotypes: T1DE1, T1DE-Int and T1DE2.

|  | T1DE1 | T1DEI | T1DE2 | pAdj<br>T1DE1<br>vs<br>T1DE2 | pAdj<br>T1DE1<br>vs<br>T1DEInt | pAdj<br>T1DEInt<br>vs<br>T1DE2 |
| --- | --- | --- | --- | --- | --- | --- |
| Age stratification | <7y | 7-12y | ≥13y |  |  |  |
| No. of T1DM | 18 | 59 | 38 |  |  |  |
| Age (years) | 4.44 ± 1.62 [18] | 12.39 ± 7.99 [59] | 20.45 ± 8.32 [38] | <0.0001 | <0.0001 | <0.0001 |
| Sex (Female/Male) | 11F/7M | 29F/30M | 18F/20M | n.s. | n.s. | n.s. |
| BMI (Kg/m<sup>2</sup>) | 15.82 ± 1.83 [18] | 17.67 ± 2.75 [59] | 21.64 ± 3.91 [38] | <0.0001 | 0.049 | <0.0001 |
| BMI_SDS | 0.14 ± 1.18 [18] | 0.16 ± 1.01 [59] | 0.5 ± 1.19 [38] | n.s. | n.s. | n.s. |
| Disease duration (weeks) | 3.53 ± 1.79 [18] | 4.02 ± 1.60 [59] | 4.42 ± 1.19 [38] | n.s. | n.s. | n.s. |
| Fasting C-peptide (pmol/L) | 158 ± 112.71 [18] | 245.86 ± 160.64 [59] | 383.65 ± 247.46 [38] | 0.0003 | 0.067 | 0.006 |
| AUC C-peptide decline (V2-V4) (absolute values, pmol/L) | 194.04 ± 109.45 [7] | 341.42 ± 303.00 [43] | 191.51 ± 418.24 [31] | n.s. | 0.015 | n.s. |
| AUC C-peptide decline (V2-V4) (% decline) | 53.8% (19.6-81.2) | 47.2% (29.5-72.10) | 17.3% (-15.6-39.3) | n.s. | n.s. | 0.003 |
| IDAA1c | 9.97 ± 2.41 [17] | 11.64 ± 2.27 [57] | 11.62 ± 2.40 [36] | 0.046 | 0.028 | n.s. |
| Insulin dose (units/kg/day) | 0.47 ± 0.31 [18] | 0.52 ± 0.25 [59] | 0.55 ± 0.28 [38] | n.s. | n.s. | n.s. |
| IAA (% positive) | 77.78 [18] | 57.62 [59] | 63.16 [38] | n.s. | n.s. | n.s. |
| IA2A (% positive) | 83.33 [18] | 77.97 [59] | 60.53 [38] | n.s. | n.s. | n.s. |
| GADA (% positive) | 61.11 [18] | 74.57 [59] | 89.47 [38] | n.s. | n.s. | n.s. |
| ZnT8A (% positive) | 38.89 [18] | 72.88 [59] | 68.42 [38] | n.s. | 0.046 | n.s. |

Number of individuals with available clinical parameters are reported in brackets. For AUC C-peptide decline (V2-V4) (% decline) values are reported as median with InterQuartile range (IQR). For AUC C-peptide decline (V2-V4) (% decline) values are reported as median with InterQuartile Range (IQR). FDR adjusted p values are reported if pAdj<0.1; N.S. is reported if pAdj>0.1.

### Clinical characterization of T1DE1 and T1DE2 endotypes in the European INNODIA cohorts

Firstly, we evaluated clinical differences across T1DE1, T1DEInt and T1DE2 endotypes.

The age-at-onset distribution for the first and second INNODIA cohorts showed high overlapping results and balanced composition **(Figure 2A-C).**

**Figure 2.**
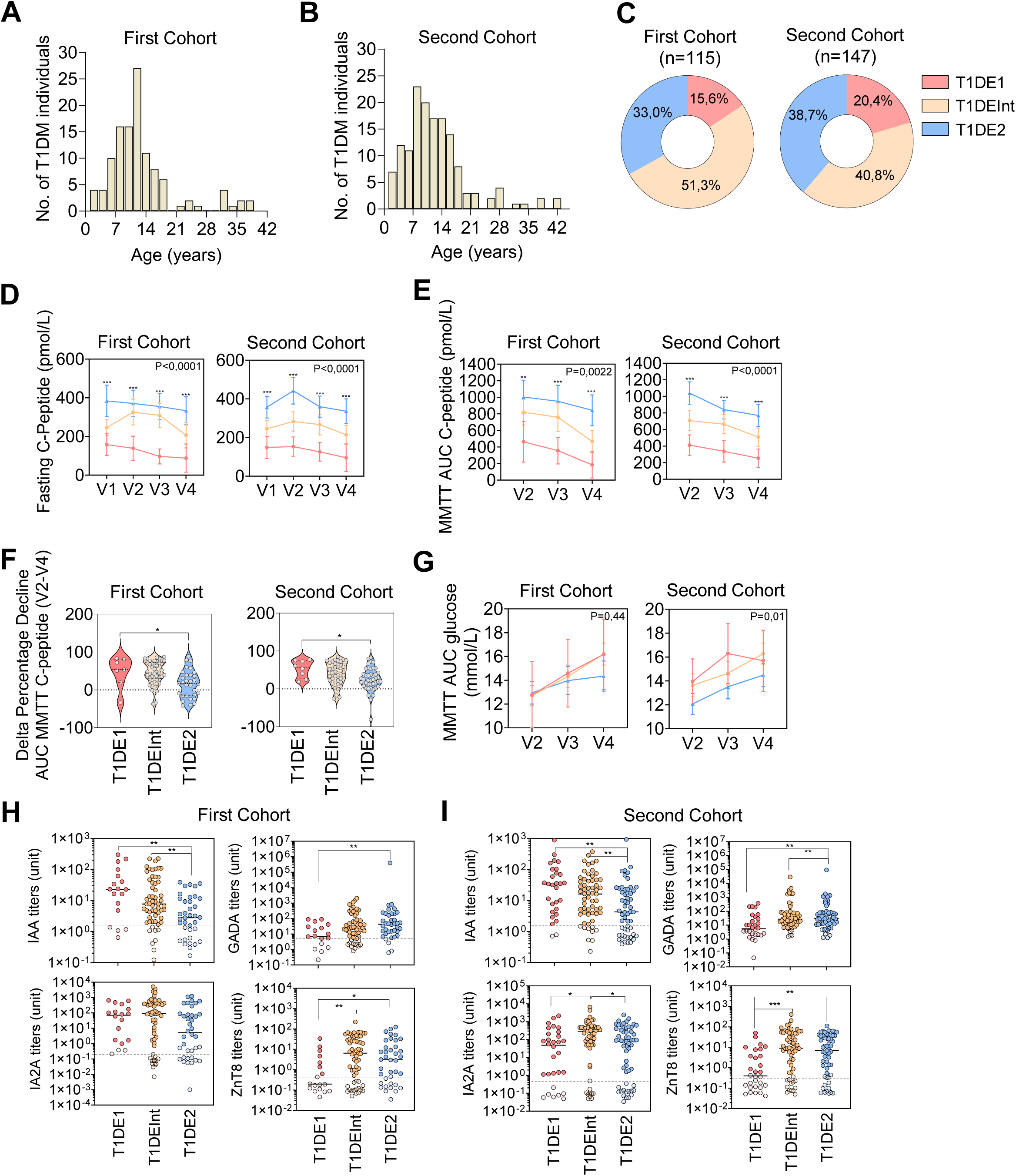
Stage 3 T1D endotypes T1DE1 and T1DE2 frequency distribution and clinical characteristic in the first and second INNODIA cohorts. (**A–B**) Age distribution (years) of individuals in the first cohort (**A**) and second cohort (**B**) at study entry, shown as histograms (y-axis: number of T1D individuals per age bin); bin width = 2y. (**C**) Endotype composition of each cohort displayed as donut charts (first cohort, n = 115; second cohort, n = 147). Participants are classified into three endotype groups (T1DE1, T1DE-Int, T1DE2; color-coded throughout as red, orange, and blue, respectively); percentages indicate the fraction of individuals assigned to each group in the corresponding cohort. (**D–F**) Longitudinal trajectories (mean ± 95%CI) by endotype: (**D**) Longitudinal fasting C-peptide concentrations (pmol/L) across study visits V1–V4 for each endotype in the first and second cohorts. (**E**) Mixed-meal tolerance test (MMTT) area-under-the-curve (AUC) C-peptide values (as labelled) across visits V2–V4 in the two cohorts. (**F**) Percentage change in MMTT AUC C-peptide between V2 and V4 V4 (higher values correspond to higher loss of β-cell function over time), shown as violin plots for each endotype (points represent individual participants; positive percentage values indicate the rate of decline in C-peptide between V” and V4, while negative value indicates a gain of β cell function; statistics using Kruskal-Wallis with Dunn’s multiple comparison test (all vs T1DE1). (**G**) MMTT AUC glucose (mmol/L) across visits V2–V4 for each endotype in both cohorts. For D, E and G, statistics were computed using Mixed-Effect model with the Geisser-Greenhouse correction; P values indicate the overall significance of the Mixed-Effect model related to the endotypes; symbols (*) indicate the significant comparison between T1DE1 (red line) and T1DE2 (blue line) at each visit (V1 – V4) using Tukey’s multiple comparison test (*P < 0.05; ** P < 0.01; ***P < 0.0001). (**H–I**) Autoantibody titers (units; log scale) for insulin autoantibodies (IAA), glutamic acid decarboxylase autoantibodies (GADA), IA-2 autoantibodies (IA2A), and ZnT8 autoantibodies (ZnT8A) in the first cohort (H) and second cohort (I), stratified by endotype; each dot represents one participant and brackets denote the pairwise comparisons highlighted in the figure. Statistics using Kruskal-Wallis with Dunn’s multiple comparison Test; significance is reported as shown in the panels (P values: * P<0.05, ** P<0.01, *** P<0.0001). “V” indicates study visits (V1: baseline; V2–V4: follow-up visits)

In the first cohort we identified n=18 T1D people assigned as T1DE1 (i.e <7 years-old), n=38 assigned to T1DE2 (i.e. ≥13 years-old) and n=59 to the intermediate endotype (T1DEInt) (i.e. between 7-12 years-old), corresponding to 15.6 %, 33.0 % and 51.3 % of the first cohort total population, respectively (**Figure 2C**, *left panel*) (**Table 1**). In the second cohort we identified n=30 T1D people as T1DE1, n=60 as T1DE2, and n=57 to the intermediate endotype, corresponding to 20.4 %, 38.7 % and 40.8 % of the second cohort total population, respectively (**Figure 2C**, right panel) (**Table 2**). Overall, the fraction of T1D individuals belonging to each endotype was highly overlapping between cohorts (**Figure 2C**).

**Table 2.**
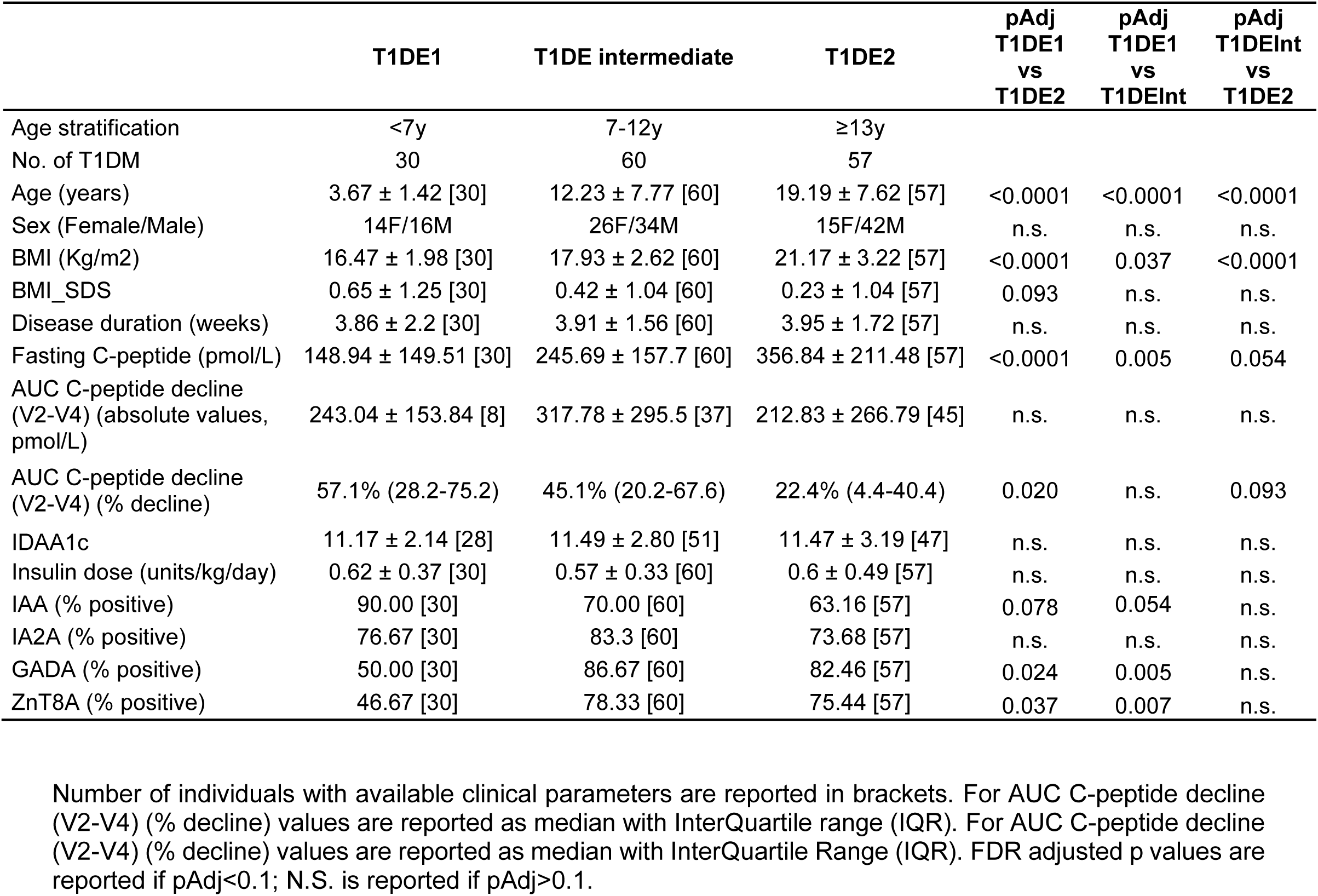
Demographics and clinical characteristics at Visit 1 (V1 - baseline) of Stage 3 T1D individuals of the INNODIA second cohort (n=147) subdivided into endotypes: T1DE1, T1DE-I and T1DE2.

As expected, people in the T1DE1 group displayed significantly lower fasting C-peptide at all visits (V1, V2, V3, V4) (**Figure 2D**) and lower MMTT AUC C-peptide at V2, V3 and V4 (**Figure 2E**) vs T1DE2 in both cohorts (linear mixed-effects model with the Geisser-Greenhouse correction, P < 0.05). In addition, the percentage of C-peptide decline (delta percentage) from V2 (3 months) to V4 (12 months after onset), corresponding to the first year of the disease, was 53.8% (n = 7, IQR: 19.6 - 81.20) for T1DE1, 47.2%(n = 43, IQR: 29.5 - 72.1) for T1DEInt and 17.3% (n = 31, IQR:-15.6 – 39.3) for T1DE2 in the first cohort (Kruskal–Wallis test, P = 0.0017), and 57.1% (n = 8, IQR: 28.2 - 75.2) for T1DE1, 45.1%(n = 37, IQR: 20.2 – 67.6) for T1DEInt and 22.4% (n = 45, IQR: 4.4 – 40.4) for T1DE2 in the second cohort (Kruskal–Wallis test, P = 0.0062) (**Figure 2F**)(**Table1 and 2**). Overall, these results showed a lower β-cell function in T1DE1 compared to T1DE2. In contrast, MMTT glucose AUC values were highly variable across endotypes and T1D people and did not mirror the decline in β cell function over the first year. Nevertheless, T1DE2 consistently showed the lowest glucose AUC across endotypes, and the linear mixed-effects model reached statistical significance only in the second cohort (**Figure 2G**). In all such analyses, similarly to other reports, when grouped using only a single parameter, T1DEInt people showed intermediate values, positioned between T1DE1 and T1DE2, though with highly variable results.

Regarding the immunological background, baseline islet autoantibody positivity and titers also differed between endotypes in both cohorts (**Figure 2H and 2I**). Notably, IAA frequency and titers were higher in T1DE1, whereas GADA and ZnT8 were lower in comparison to T1DE2 (P < 0.05, Kruskal-Wallis with Dunn’s multiple comparison test); IA-2A showed less pronounced differences in both cohorts (**Figure 2H and 2I**, lower panels) (**Table 1 and Table 2**).

Collectively, these patterns, reproduced across two independent but overlapping T1D cohorts, support lower β-cell function during the first year after diagnosis and a higher burden of immunological markers in T1DE1, thereby reinforcing and extending the clinical characterization of these endotypes as reported in previous studies (4)(31–33)(34)(35)(36)(37).

### Plasma miR-150-5p and miR-375-3p are increased in T1DE1 at diagnosis

To profile circulating miRNAs distinguishing T1DE1 from T1DE2, we re-analyzed small RNA-seq datasets obtained from plasma samples of T1D individuals of the first and second INNODIA cohort. Such analyses were conducted separately in the two cohorts with a specific focus on those miRNAs differentially expressed and shared between them.

Differential expression analysis revealed 29 miRNAs upregulated with no downregulated miRNAs in T1DE1 versus T1DE2 in the first cohort whereas in the second cohort we observed 45 downregulated and 3 upregulated miRNAs (P < 0.05, FDR-adjusted) (**Supplementary Figure 1A and 1B**). Notably, two miRNAs, miR-150-5p and miR-375-3p, were concordantly higher in plasma of T1D people belonging to T1DE1 in comparison to T1DE2 across both cohorts (**Figure 3A) (Supplemental Tables 2A and 2B).** This was evident in small RNA seq-derived normalized reads counts plots (**Figure 3A**) and independently validated by using digital droplet PCR (ddPCR) analysis, which confirmed higher copies/µL of both miRNAs in T1DE1 in comparison to T1DE2 in the first and second cohort (P< 0.05, Kruskal-Wallis with Dunn’s multiple comparison test) (**Figure 3B**).

**Figure 3.**
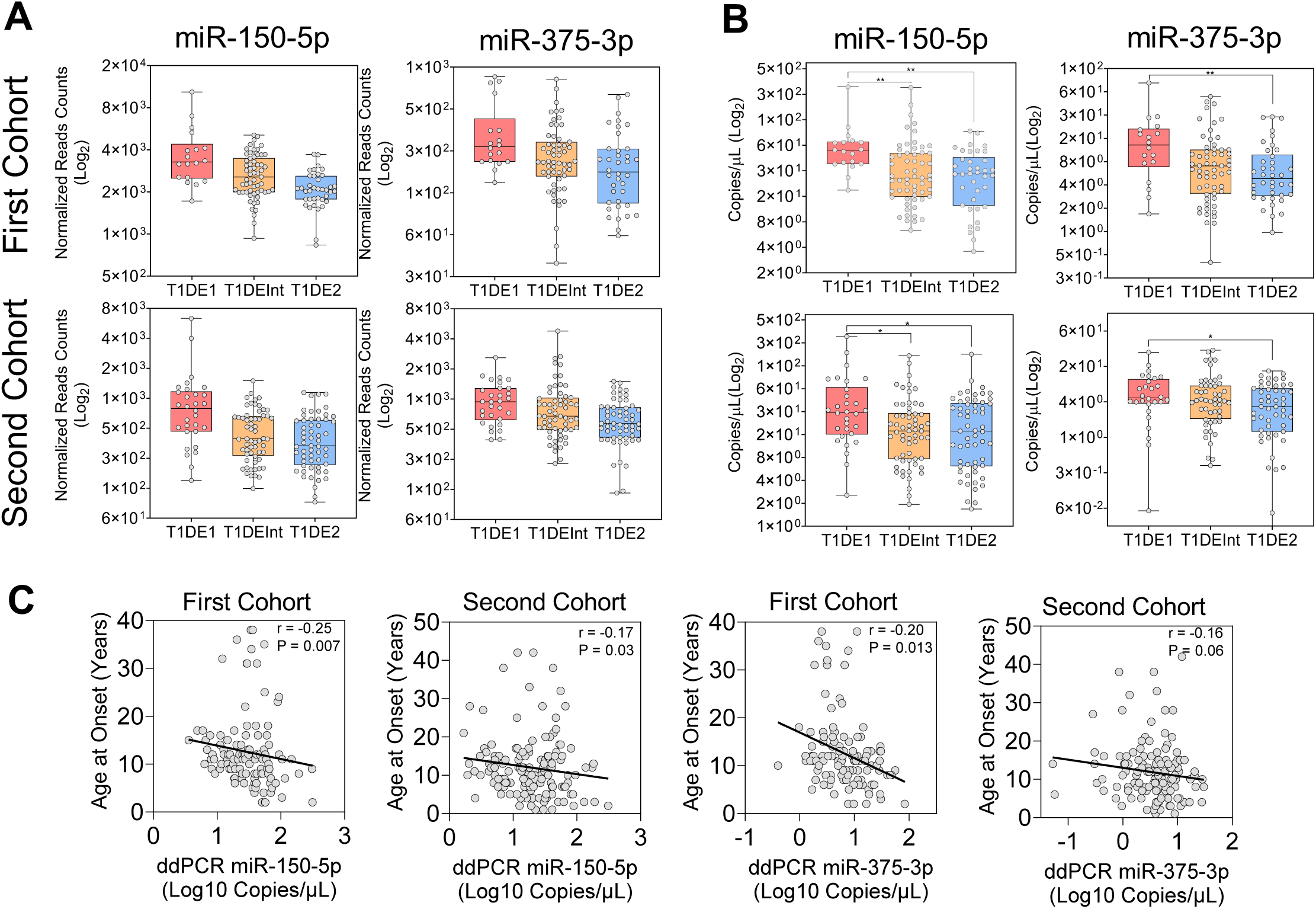
miR-150-5p and miR-375-3p distinguish T1DE1 and T1DE2 endotypes. **(A)** Small RNA-seq quantification of circulating miR-150-5p and miR-375-3p shown as normalized read counts (log2) across endotypes (T1DE1, T1DEI, T1DE2) in the first (top) and second (bottom) cohorts. **(B)** Independent validation by ddPCR, reported as absolute concentration (copies/µL; log2 scale), across the same endotype groups in each cohort. Boxplots show median and interquartile range (IQR), whiskers indicate 1.5×IQR; statistics using Kruskal-Wallis with Dunn’s multiple comparison test; group-wise significance is indicated by brackets and asterisks (*p<0.05, **p<0.01, ***p<0.001). **(C)** Scatter plots of ddPCR levels (log10 copies/µL) versus age at onset in the first and second cohorts; Spearman correlation coefficients and two-sided p values are reported in each panel; dots represent individual participants. (**D**) Receiver operating characteristic (ROC) curves evaluating the ability of circulating miR-150-5p and miR-375-3p to discriminate T1DE1 vs T1DE2 in the first and second cohorts. ddPCR copies/ µL were used to compute the ROC analysis. Each point represents individual value. The diagonal dashed line indicates performance expected by chance (AUC = 0.5).

As expected, both miR-150-5p and miR-375-3p levels, as measured using ddPCR, were correlated inversely with age-at-onset both in the first and second cohort, showing significant though low Spearman *rho* values ([miR-150-5p: first cohort r=-0.25 – P = 0.007; second cohort r=-0.17 – P = 0.03. miR-375-3p: first cohort r=-0.20 – P = 0.013; second cohort r=-0.16 – P = 0.06) (**Figure 3C**) (**Supplemental Figure 1C-1D**) (**Supplementary Table 3**). These results suggest that although miR-150-5p and miR-375-3p display endotype-associated differences, their correlations with age at diagnosis were modest, suggesting that these miRNAs are not merely a linear readout of age at T1D onset.

However, because T1DE1 and T1DE2 are intrinsically age-linked, we performed two complementary analyses: (*i*) we tested whether miR-150-5p and miR-375-3p are associated with age in age-matched, publicly available pediatric/adolescents circulating small RNA-seq datasets; and (*ii*) we applied age-adjusted models in the INNODIA T1D cohorts to separate age effects from endotype-associated biology.

In two independent pediatric serum small RNA-seq datasets - PRJNA793584 (healthy controls and coeliac disease) (38) and GSE134897 (childhood asthma) (39), both spanning an age range comparable to our T1D cohorts (**Table 3**) - miR-150-5p and miR-375-3p showed no association with age when correlating normalized reads counts with age in controls, coeliac disease, or asthma groups (**Supplemental Figure 2A–F**) (**Supplemental Table 3**).

**Table 3.**
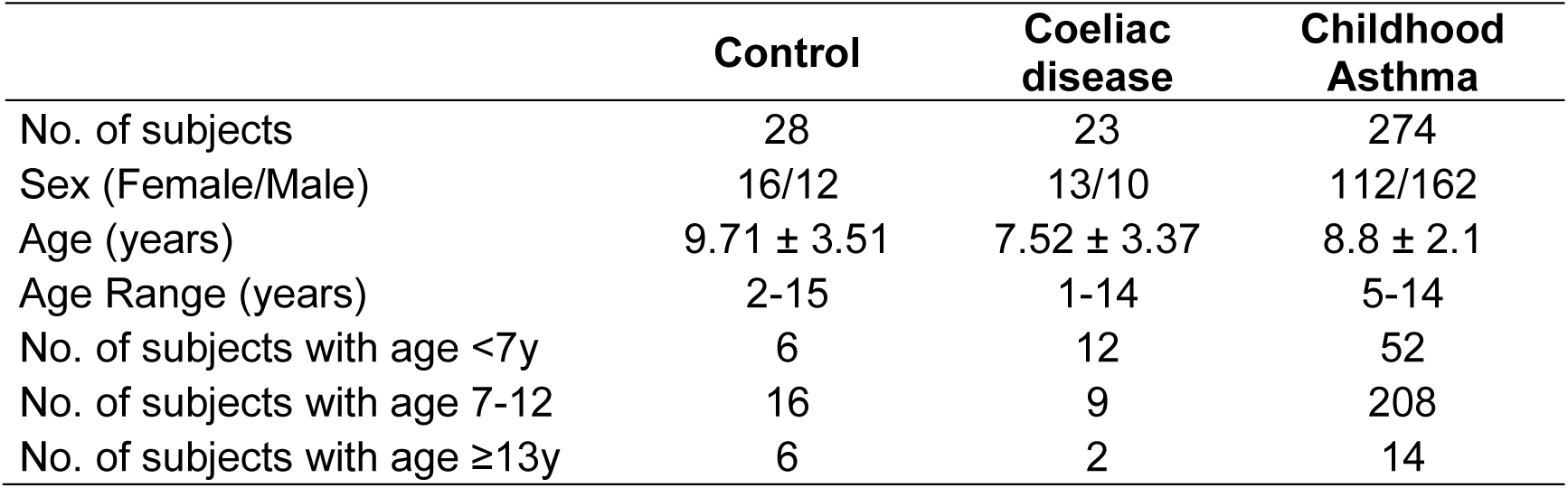
Demographics and main characteristics of non-diabetic cohorts.

|  | **Control** | **Coeliac disease** | **Childhood Asthma** |
| --- | --- | --- | --- |
| No. of subjects | 28 | 23 | 274 |
| Sex (Female/Male) | 16/12 | 13/10 | 112/162 |
| Age (years) | 9.71 ± 3.51 | 7.52 ± 3.37 | 8.8 ± 2.1 |
| Age Range (years) | 2-15 | 1-14 | 5-14 |
| No. of subjects with age <7y | 6 | 12 | 52 |
| No. of subjects with age 7-12 | 16 | 9 | 208 |
| No. of subjects with age ≥13y | 6 | 2 | 14 |

Although miR-375 and miR-150-5p did not correlate with age in non-T1D paediatric/adolescent cohorts, we further tested whether the miRNA-endotype association observed in INNODIA T1D cohorts could reflect residual age effects by repeating the differential expression analyses with age included as a covariate in the statistical model. As summarized in **Table 4**, miR-375 remained significantly upregulated in T1DE1 versus T1DE2 only in the second cohort, whereas the association was attenuated and no longer significant in the age-adjusted analysis of the first cohort (P = 0.33). Importantly, miR-150-5p remained robustly upregulated in T1DE1 versus T1DE2 both in the first and second cohort even after age adjustment (P=0.02 and P=1.47×10^-7^, respectively). Collectively, these results indicate that, despite the intrinsic link between age and the T1D endotypes, miR-150-5p and, less strongly, miR-375 showed a reproducible association that cannot be attributed to age alone, thus suggesting these molecules as potential markers of the underlying biology of the two endotypes.

**Table 4.**
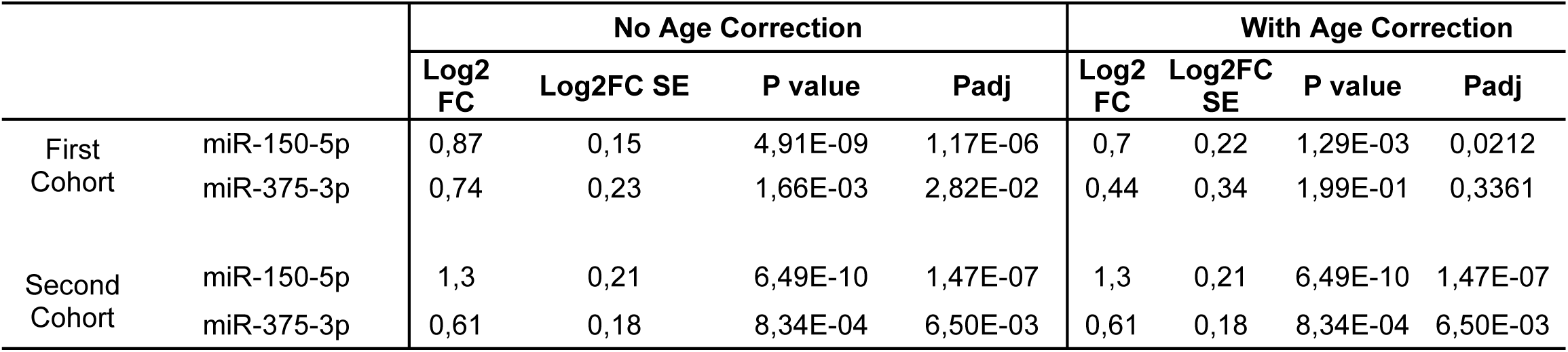
Results of the differential expression analysis on circulating miRNAs in T1DE1 vs T1DE2, with and without age correction.

### miR-150-5p is highly expressed in B-and T-lymphocytes but its circulating levels are not associated with frequencies of relevant peripheral blood circulating immune cell subsets

To gain insights into the putative cellular origin of circulating miR-375-3p and miR-150-5p, we interrogated a large curated dataset of small RNA-seq experiments (isomiRdb) (40) to investigate whether their expression could be enriched in a specific cell type across a wide range of different human tissues and cell types collection.

As expected, miR-375-3p is highly expressed and enriched in pancreatic islets and β cells (data not shown). These results are confirmed by multiple previous studies showing its expression and relevance in islet and β cell context (41–44).

Regarding miR-150-5p levels across human tissues, we observed that it is predominantly expressed in blood and plasma (**Supplementary Figure 3A**). Interestingly, across specific human cell types, miR-150-5p is highly expressed in B-, CD8^⁺^ and CD4^⁺^ T lymphocytes as well as Natural Killer cells (**Figure 4A and Supplementary Figure 3B**). These findings corroborate previous studies which highlighted the high expression and important functional role(s) for miR-150-5p in B-lymphocytes (45–47), as well as T-lymphocytes (48–50).

**Figure 4.**
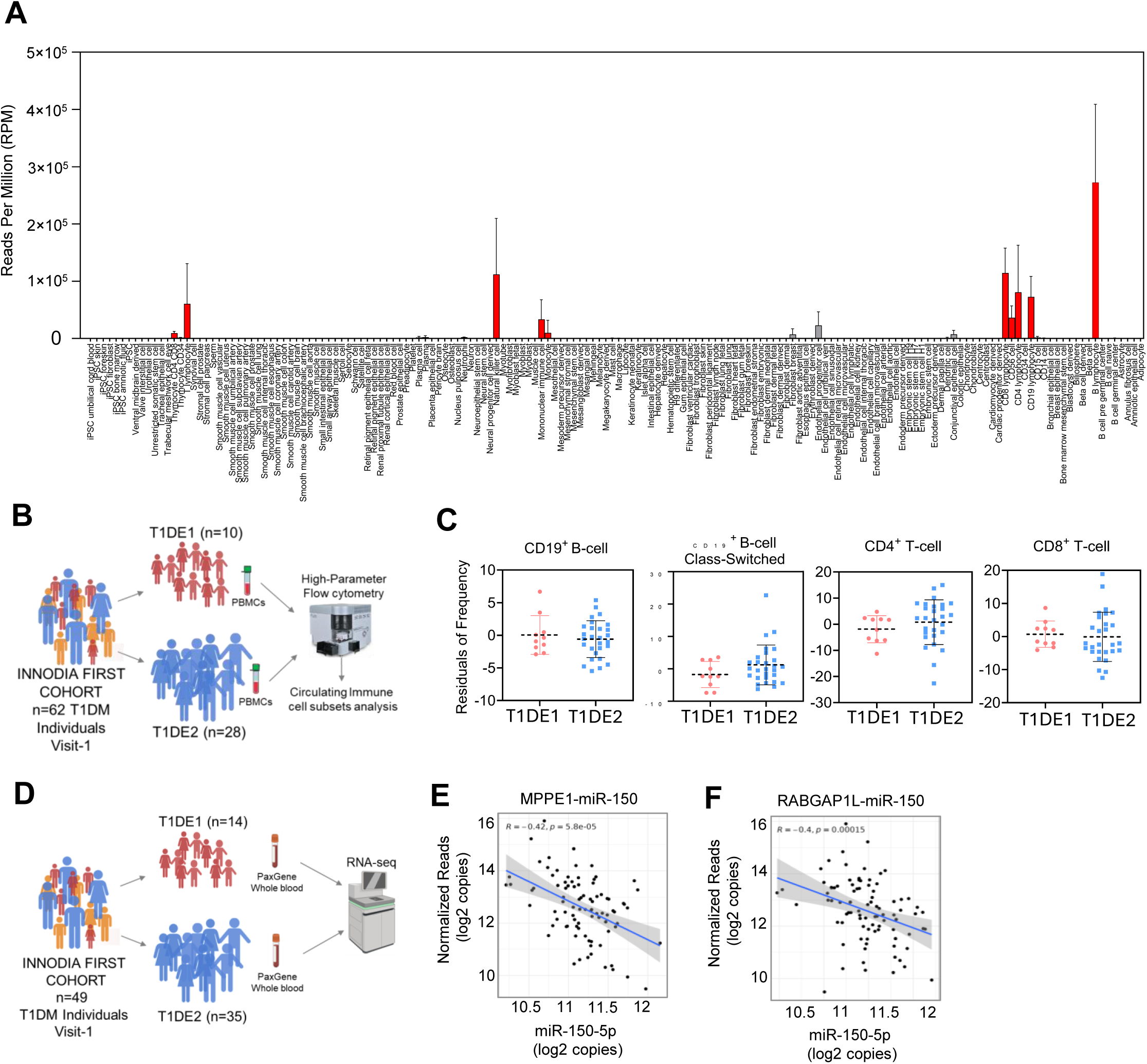
miR-150-5p is highly expressed in immune cells and is inversely correlated with immune-associated genes. (**A**) Reference expression of the endotype-associated miRNA miR-150-5p across human tissues and primary cell types (n=100) from a public miRNA atlas (isomiRDB), reported as reads per million (RPM). Expression is strongly enriched in hematopoietic/lymphoid compartments (highlighted in red), consistent with a predominant immune-cell origin. Bars show mean ± S.D. (**B**) Study schematic: PBMCs collected at visit 1 from the INNODIA First Cohort (n = 62) were stratified by endotype (T1DE1, n = 10; T1DE2, n = 28) and analyzed by high-parameter flow cytometry to quantify circulating immune-cell subsets. (**C**) Comparison of selected lymphocyte populations between endotypes, shown as residuals of cell frequency (i.e., covariate-adjusted values): CD19^⁺^ B cells, class-switched CD19^⁺^ B cells, CD4^⁺^ T cells, and CD8^⁺^ T cells. Each dot represents one individual (T1DE1 in red; T1DE2 in blue); horizontal lines and whiskers summarize group central tendency and dispersion. (**D**) Whole-blood transcriptomics study design and number of T1D people belonging to INNODIA first cohort with available small RNA-seq analysis subdivided into T1DE1 and T1DE2 endotypes. (**E-F**) Correlation between miR-150-5p (x axis) and (E) MPPE1 and (F) RABGAP1L reported as normalized reads (log2+1). Each dot represents a T1D individual. Spearman r and P values are reported. Correlation line (in blue) with confidence interval (gray zone) is reported for each gene.

Therefore, given the reported enrichment of miR-150-5p in both T and B lymphocytes, we investigated whether its higher plasma levels in T1DE1 in comparison to T1DE2 could simply reflect a greater circulating abundance of these cell types in T1DE1. To test this, we re-analysed immunomic datasets obtained by high-parameter flow cytometry in a subset of the INNODIA first cohort (67/115 individuals), including n = 10 T1DE1 and n = 28 T1DE2 (**Figure 4B**). Contrary to this expectation, we detected no differences between endotypes in the frequencies of CD19^⁺^ B cells, class-switched CD19^⁺^ B cells, CD4^⁺^ T cells or CD8^⁺^ T cells (**Figure 4A**). These data argue against a peripheral blood cell-composition explanation for the increased circulating miR-150-5p levels but, instead, point toward qualitative differences in lymphocyte state, putatively in the target organ (i.e. insulitic infiltrate).

### The expression of miR-150-5p is negatively correlated with immune-associated genes MPPE1 and RABGAP1L

As part of the INNODIA consortium, we had access to whole-blood transcriptomic data from a subset of newly diagnosed individuals in the first cohort. Particularly, whole-blood RNA-seq profiling at visit 1 was available for 49 participants, including 14 classified as T1DE1 and 35 as T1DE2 (**Figure 4D**). To explore the biological relevance of circulating miR-150-5p and its potential effect(s) on the transcriptome of circulating immune cells, we correlated its plasma levels, measured with ddPCR, with whole-blood gene expression. This analysis identified two transcripts showing significant inverse associations with miR-150-5p: MPPE1 (Spearman’s ρ = −0.42, P = 5.8×10^⁻⁵^, FDR-adjusted P = 0.03; **Figure 4E**) and RABGAP1L (Spearman’s ρ = −0.40, P = 1.4×10^⁻⁴^, FDR-adjusted P = 0.04; **Figure 4F**). Notably, both genes are reported as predicted target of miR-150-5p, according to TargetScan7.2 analysis (**Supplemental Table 4**). These findings support MPPE1 and RABGAP1L as miR-150-5p–linked transcripts in vivo and suggest that higher circulating miR-150-5p is associated with a coordinated shift in whole-blood gene expression that may contribute to the molecular differences between T1D endotypes.

To contextualize these associations at the cellular level, we examined the expression patterns of MPPE1 and RABGAP1L using the online tool Human Protein Tissue Atlas (https://www.proteinatlas.org/). Both genes were preferentially expressed in immune cell populations; particularly, MPPE1 showed enrichment in neutrophils and natural killer (NK) cells, whereas RABGAP1L was predominantly enriched in non-classical monocytes and neutrophils. Given the autoimmune nature of T1D, the involvement of these innate and cytotoxic immune subsets provides biological plausibility to the observed correlations and supports the hypothesis that secreted/circulating miR-150-5p is associated with molecular programs embedded within specific immune compartments that are implicated in T1D pathogenesis.

### Combined miR-150-5p and miR-375-3p re-stratify T1D individuals in two clinically-distinct groups: miRC1 and miRC2

We observed that both miR-150-5p and miR-375-3p showed similar discriminatory capacity in classifying T1D individuals as T1DE1 or T1DE2, as calculated by ROC curves analysis; these results are consistent across both INNODIA T1D cohorts, albeit with a lower performance in the second one (**Figure 5A**). In the first cohort, circulating miR-150-5p significantly discriminates the two endotypes with an AUC of 0.78 (specificity:65.7%; sensitivity: 88.8%; p=0.0006), while miR-375-5p showed an AUC of 0.72 (specificity:68.4%; sensitivity: 77.7%; p=0.0005). In the second cohort, both miRNAs showed a lower AUC (miR-150-5p: 0.65; miR-375-3p: 0.63) and lower specificity and sensitivity values. Nonetheless, in both cohorts, the two circulating miRNAs were able to discriminate T1DE1 and T1DE2 with statistically significant results (**Figure 5A**).

**Figure 5.**
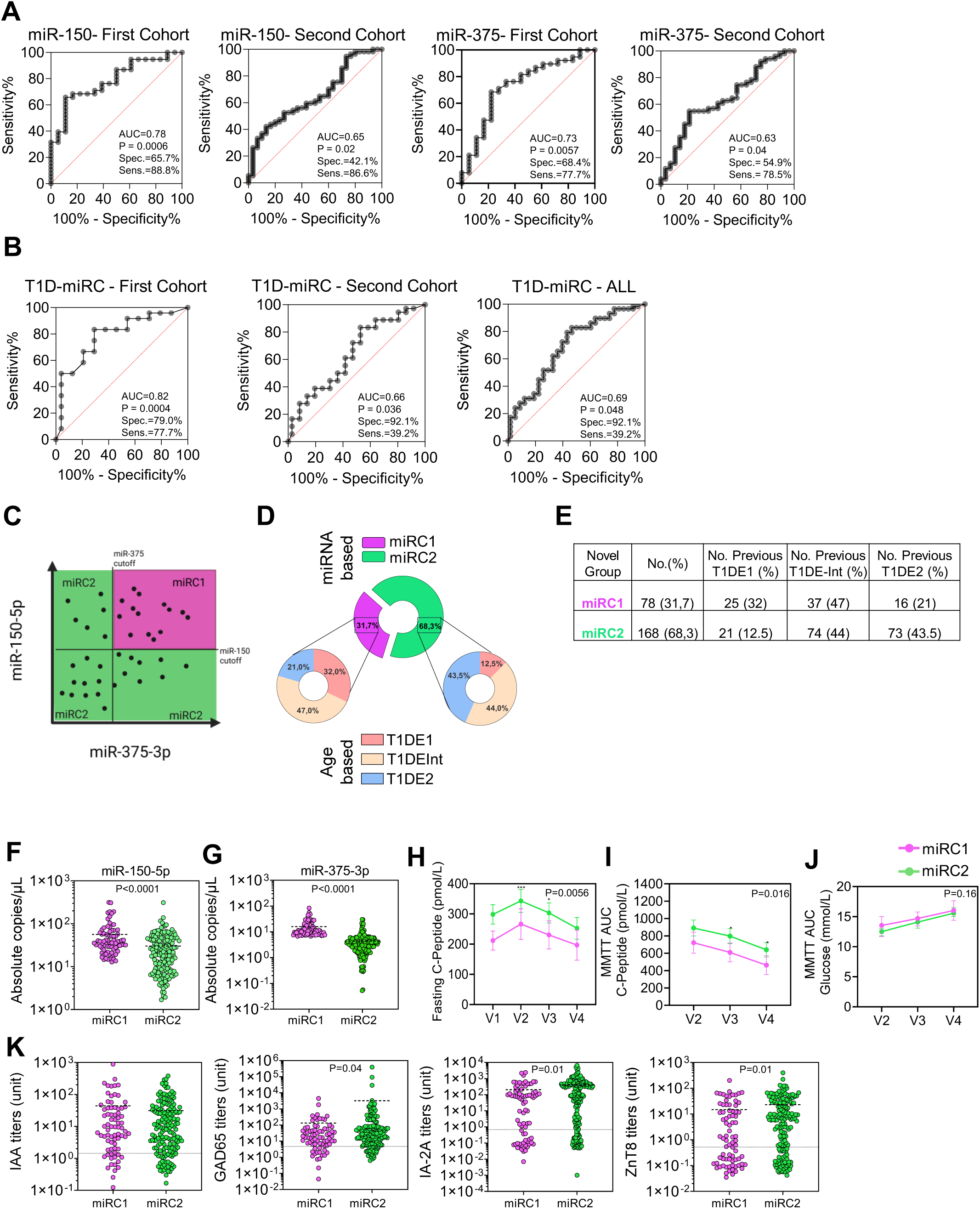
Performance of combined circulating miRNAs for T1DE re-classification and association of miRC subgroups to clinical characteristics. (**A**) Receiver operating characteristic (ROC) curves for miR-150 and miR-375 evaluated separately in the first and second cohorts; AUC, P value, specificity and sensitivity are reported in each panel. (**B**) ROC performance of the combined logistic regression miRNA classifier (T1D-miRC) in the first cohort, second cohort, and in the pooled dataset (“ALL”). (**C**) Schematic overview of the two classifications using a logical classifier: miRC1=both miRNA > cutoff thresholds; miRC2= all other cases. (**D**) Composition of miRNA-based endotypes (miRC1 and miRC2) and their correspondence with previously defined T1D endotypes (T1DE1, T1DEInt, T1DE2) shown as proportional donut charts. (**E**) Contingency table reporting sample size and the distribution (n, %) of previous endotypes within each miRNA-defined endotype. (**F, G**) Circulating absolute levels (copies/mL) of miR-150-5p (**F**) and miR-375-3p (**G**) in miRC1 vs miRC2; each dot represents one individual. (**H**) Longitudinal fasting C-peptide across visits (V1–V4) stratified by miRC endotype (group trajectories shown with error bars as plotted). (**I, J**) MMTT AUC for C-peptide (**H**) and glucose (**I**) across visits (V2–V4) by miRC endotype; exact P values are indicated for linear mixed-effect model with Tukey’s multiple comparison test for miRC1 vs miRC2 at each visit: *p<0.05; **p<0.01; ***p<0.001. (**K**) Autoantibody titers (IAA, GAD65, IA-2A, ZnT8; log scale) in miRC1 vs miRC2; dashed lines denote assay cut-offs, with P values reported where significant (Mann Whitney U test).

However, it remains unclear whether these miRNAs can also reliably classify T1DInt individuals into different groups showing distinct clinical and phenotypical characteristics potentially resembling T1DE1 or T1DE2 endotypes. Hence, given the distinct roles of miR-150-5p and miR-375-3p in capturing different sides of the pathogenetic mechanism in T1D (immune cells activation - β cells stress and/or destruction) we explored their potential combination to re-classify T1D individuals, including T1DInt ones, into clinically distinct subgroups. For this purpose, we employed two classification strategies. First, we quantified the discriminatory ability of combined miR-150-5p and miR-375-3p using a multivariable logistic regression score and ROC/AUC analyses on extreme endotypes (T1DE1 vs T1DE2). Second, for downstream clinical stratification, we adopted an interpretable rule-based signature (“AND” classifier) based on miRNA-specific ROC curves Youden-derived ddPCR expression cutoffs, calculated on T1DE1 and T1DE2 individuals of combined cohorts (n=135) and applied fixed thresholds to the full population with available ddPCR miRNAs values (n=246), including T1DInt subjects.

ROC/AUC curve analyses of combined miRNAs (miRCombo-miRC) on T1DE1 and T1DE2 endotypes showed only marginal improvements compared to the analysis of the single miRNAs and only for some of the ROC parameters, even though all reached statistical significance, including the ROC curve analysis of all individuals from both cohorts (**Figure 5B**). Hence, given the lack of dramatic improvement when using this combination analysis, we decided to apply a logical “AND” rule, whereby subjects were classified as miRNA-high (miRC1) only if both miR-150-5p and miR-375-3p satisfied their respective Youden-derived cutoffs, while all remaining subjects were classified as miRNA-low (miRC2) (**Figure 5C**). Using this classification, n=78 T1D individuals are included in the miRC1 while n=168 in the miRC2 subgroup (**Figure 5D and 5E**). These novel subgroups include individuals that were previously assigned to one of the two endotypes (T1DE1, T1DE2) or, importantly, to the intermediate subgroup (T1Dint). Interestingly, in this novel re-classification, previous T1DInt (n=111/246) are almost equally distributed into the two subgroups (miRC1: 47% of previous T1DInt; miRC2: 44% of previous T1Dint). The proportion of T1DE1 individuals defined as miRC1 was 32%, while the proportion of T1DE2 ones defined as miRC2 was 43.5% (**Figure 5D**). As expected, T1DE1 individuals were more prevalent in miRC1 (32% in miRC1 vs 12.5% in miRC2) while T1DE2 individuals were more prevalent in miRC2 (43.5% in miRC1 vs 21% in miRC2) (**Figure 5D and 5E**). Of interest, using this classifier, several T1DE1 and T1DE2 individuals are re-assigned to one of these novel groups, independently of the age at onset.

As expected, both miRNAs are strongly increased in miRC1 compared to miRC2 (**Figure 5F and 5G**). More importantly, miRC1 individuals showed significantly lower fasting C-peptide (linear mixed-effects model, p=0.0056) (**Figure 5H**) and lower MMTT AUC C-peptide at V2, V3 and V4 (mixed-effects model, p=0.016), while MMTT AUC glucose did not differ between miRC1 and miRC2, similar to earlier data for T1DEs. Additionally, islet autoantibody profiles showed increased titers of GAD65A, IA-2A and ZnT8A, while no differences are observed for IAA.

As expected, and as in line with previously described age endotypes, miRC1 and miRC2 individuals showed significantly different age at onset (miRC1: 9.87±7.16y - miRC2: 13.23±7.67; p=0.0004 Mann Whitey U test), as well as BMI and BMI-SDS (**Table 5**).

**Table 5.** Demographics and clinical characteristics at Visit 1 (V1 - baseline) of stage 3 T1D individuals of the combined INNODIA first and second cohort (n=246) subdivided into miRC1 and miRC2.

|  | miRC1 | miRC2 | P value |
| --- | --- | --- | --- |
| No, of T1DM | 78 | 168 |  |
| Age (years) | 9,87 ± 7,16 [78] | 13,23 ± 7,67 [168] | 0,0004 |
| Sex (Female/Male) | 38F/40M | 68F/100M | n.s. |
| BMI (Kg/m<sup>2</sup>) | 17,38 ± 2,59 [78] | 19,41 ± 3,69 [168] | 0,0006 |
| BMI_SDS | 0,12 ± 1,03 [78] | 0,43 ± 1,12 [168] | n.s. |
| Fasting C-peptide (pmol/L) | 211,88 ± 136,86 [76] | 298,50 ± 209,27 [168] | 0,0097 |
| IDAA1c | 11,51 ± 2,56 [74] | 11,34 ± 2,65 [162] | n.s. |
| Insulin dose (units/kg/day) | 0,56 ± 0,29 [78] | 0,55 ± 0,38 [164] | n.s. |
| AUC C-peptide decline (V2-V4)<br>(absolute values, pmol/L) | 233,76 ± 232,31 [n=42] | 284,00 ± 326,51 [n=120] | n.s.<br>n.s. |
| AUC C-peptide decline (V2-V4)<br>(% decline) | 38,23 ± 32,81 [n=42] | 32,37 ± 36,11 [n=120] | n.s.<br>n.s. |
| IAA (% positive) | 76,92 [78] | 79,17 [168] | n.s. |
| IA2A (% positive) | 69,23 [78] | 79,17 [168] | n.s. |
| GADA (% positive) | 75,64 [78] | 80,36 [168] | n.s. |
| ZnT8A (% positive) | 60,26 [78] | 73,81 [168] | n.s. |

Number of individuals with available clinical parameters are reported in brackets. For AUC C-peptide decline (V2-V4) (% decline) values are reported as median with InterQuartile range (IQR). For AUC C-peptide decline (V2-V4) (% decline) values are reported as median with InterQuartile Range (IQR). FDR adjusted p values are reported if pAdj<0.1; N.S. is reported if pAdj>0.1.

Collectively, these data show that miR-150-5p and miR-375-3p can be combined to classify T1D individuals at onset into two distinct subgroups (miRC1 and miRC2) which partially resemble pivotal clinical characteristics of T1DE1 and T1DE2 and can also be used to assign previously undefined T1DInt subjects to one of these two subgroups, independently of age at disease onset.

## DISCUSSION

Growing evidence indicates that T1D is not a single uniform disease but a highly heterogeneous group of biologically distinct endotypes (6, 51). Seminal histopathological studies by Leete and colleagues have demonstrated for the first time that children diagnosed at an early age exhibit a highly aggressive pattern of insulitis, characterized by dense immune cell infiltration, with an abundance of CD20^⁺^ B cells (CD20-high), pronounced interferon-stimulated gene signatures, and near-complete β-cell loss at clinical onset (7)(52). More recent integrative work has expanded this view, proposing that additional immunogenetically defined endotypes may exist, shaped by interactions among age at onset, HLA haplotype, inflammatory pathways and metabolic resilience (8). A recent study showed that children progressing to T1D exhibit distinct early cellular immune responses depending on whether insulin or GAD autoantibodies appear first, consistent with endotype-specific disease trajectories (53). Together, these findings provide a robust framework in which the heterogeneity of T1D reflects distinct pathogenic pathways present prior to diagnosis, highlighting the need for non-invasive biomarkers capable of distinguishing endotypes, particularly from a pathological mechanism perspective, for use in clinical practice for the administration of different immunotherapies. Consistent with this view, a recent international expert forum underscored the marked heterogeneity of T1D and its clinical implications, and renewed the call for sustained efforts to identify additional determinants and biomarkers of disease, both at the population level and within biologically and clinically defined subgroups (51).

Within this conceptual landscape, our study firstly characterised the clinical phenotypes of T1D endotypes in the INNODIA natural history cohort (first and second cohort), and then focused on circulating miRNAs by identifying miR-150-5p and miR-375-3p as part of a highly consistent molecular signature distinguishing the early-onset, aggressive T1DE1 endotype from the more indolent T1DE2 endotype, which perfectly recapitulate and confirm the clinical signatures of these endotypes as previously observed (2). Finally, we were able to use these two miRNAs to re-stratify T1D individuals into two distinct groups which also include the T1D intermediate endotype (7-12y). The reproducibility of the miRNAs profiling results, in relation to the two miRNAs, in two independent INNODIA T1D populations, supports the notion that miR-150-5p and miR-375-3p elevations may reflect an endotype-specific immunopathology. This interpretation aligns with a rapidly expanding body of literature positioning circulating microRNAs as stable and informative markers of immune dysregulation and/or β-cell dysfunction in T1D (54)(55). Additionally, both miR-150-5p and miR-375-3p were not associated with age in non-T1D cohorts, thus suggesting that the observed differences might not be a mere effect of the age at onset; moreover, miR-150-5p, but not miR-375-3p, remained statistically significant even after correction with age-at-onset in both T1D INNODIA cohorts, thereby reinforcing this concept.

miR-375-3p and miR-150-5p are not new in the T1D panorama. miR-375-3p is highly expressed and enriched in pancreatic islets (43, 56) with a demonstrated role in β-cell insulin secretion, development/differentiation and maintenance of β-cell mass (57, 58). MiR-375-3p was found in the plasma of individuals with T1D and resulted one of the most associated circulating miRNAs to T1D (59). Notably, β-cell destruction is sufficient to cause elevations of miR-375 levels in the blood and its upregulation has been observed in plasma/serum of T1D people in comparison to controls, or during the early stages of islets transplant both in animal models and in the human context (60–66). Hence, its upregulation in plasma of T1DE1 individuals may indicate a condition of high β-cell stress and death, in line with the more severe clinical phenotype reported in this study and in other previous studies. These view is also in line with the histopathological features of T1DE1 endotype characterized by lower β-cell mass potentially due to the aggressive lymphocytic infiltrates observed in the pancreas from T1DE1 donors (67).

MiR-150-5p is highly expressed in immune cells and previous studies have pointed to its central role as a regulator of multiple immune cell functions (45),(68),(69),(46); our findings corroborate its high expression in immune cells, particularly in B-lymphocytes. In these cells, miR-150-5p can control differentiation and activation (47, 70). Of note, upon B-and T-lymphocyte activation, intracellular miR-150-5p is rapidly downregulated through its increased secretion and extracellular disposal, a mechanism adopted by lymphocytes to rapidly modulate multiple intracellular pathways. Hence, lymphocyte activation phenomena may correspond to increased extracellular and plasma circulating levels of miR-150-5p (71–73). In this framework, the upregulation of miR-150-5p in T1DE1 aligns closely with the aggressive immunopathological features of this endotype and the increased frequency of B-lymphocytes found in the pancreas of T1DE1 donors (CD20-high); these characteristics correspond also to a heightened in situ IFN signature, a common read-out of increased activity of pancreas lymphocytic infiltrates (Ref). However, miR-150-5p is not merely an intracellular regulator, given that this miRNA is secreted into extracellular vesicles by activated lymphocytes (71), making it detectable in plasma as a dynamic signal of immune activation and a communication signal. Of note, miR-150-5p is increased in human islet grafts and in the circulation during islet xenograft rejection and β-cell destruction prior to hyperglycaemia and has been suggested as an early biomarker for islet xenograft rejection or for the early stages of the disease (66). Hence, we hypothesize that its increased expression in plasma of T1DE1 individuals may reflect the ongoing activation of lymphocytes which characterizes the increased severity of the disease in this endotype. The biological plausibility of miR-150-5p as an endotype marker for T1DE1 is also reinforced by its well-established immunoregulatory functions. MiR-150-5p shapes hematopoietic lineage decisions by targeting MYB (74) and modulates T-cell differentiation and effector programs via pathways including Notch3 (75). Extensive work in both mouse and human systems shows that miR-150 controls thresholds for T-and B-cell activation, memory formation and metabolic programming (68) (45),(47),(50),(76),(70). The negative correlation we observed between circulating miR-150-5p and the expression of immune related genes such as MPPE1 and RABGAP1L, enriched in neutrophils, NK cells and non-classical monocytes, suggests broader immune-network differences between endotypes that may involve both adaptive and innate immune pathways though new mechanisms of communication between adaptive and innate immune cells. Although such relationships are correlative, they provide initial evidence that circulating miR-150-5p may contribute to, or reflect the reconfiguration of, immune communication circuits that distinguish T1DE1 from T1DE2.

Overall, our data alongside available studies suggests that these miRNAs could represent circulating markers of two different sides (immune activation, β cells destruction) of the same disease. As a results, using ddPCR absolute copies/µL values, we identified specific miRNA cutoffs, trained on a ROC curve model which incorporate exclusively T1DE1 and T1DE2 individuals of the two combined cohorts, that can be used to re-stratify all T1D individuals. Importantly, this approach allowed us to assign T1DInt subjects to one of the two defined subgroups, that we named miRC1 and miRC2.

T1D individuals in miRC1 group, characterised by the highest miRNAs expression (both miR-150 and miR-375 high), showed lower fasting and stimulated c-peptide, while in miRC2 they showed opposite characteristics (i.e. a less compromised β cell function) and higher titers or frequency of GADA, IA-2A and ZnT8. Interestingly, such re-classification assigned previously defined T1DE1 or T1DE2 individuals to different groups (miRC1 or miRC2). This result suggests that individuals with T1D can be classified into specific endotypes by approaches that are independent of the age at onset. Most importantly, T1DInt individuals can be assigned to one or other of these subgroups by this means.

Several limitations of this study should be considered. First, although the INNODIA cohorts represent among the largest resources available for circulating miRNA profiling at new-onset T1D, T1DE1 remains intrinsically under-represented because of disease epidemiology, which limits power for endotype-specific subgroup analyses. Second, immunophenotyping and transcriptomic data were available only for subsets of participants, reducing our ability to resolve rare immune populations and to capture activation or differentiation states that may be most relevant to circulating miR-150-5p signals. Third, the study is cross-sectional at diagnosis, precluding inference on predictive performance and limiting conclusions on longitudinal trajectories of miR-150-5p and miR-375-3p in relation to progression or treatment response. Fourth, associations between circulating miRNAs and immune gene-expression patterns remain correlative and will require functional validation in purified cell subsets and ideally perturbation-based approaches. Finally, because the INNODIA Master Protocol did not include recruitment of unrelated control people without diabetes for small RNA screening, we did not generate an internal control reference for absolute disease-versus-health comparisons. Importantly, it should be noted that the primary aim of this work was not to define “T1D versus control” biomarkers, but to interrogate differences between established T1D endotypes at diagnosis to gain insight into underlying immunopathology and to improve clinically meaningful stratification.

Despite these limitations, our data robustly support the concept that circulating miR-150-5p and miR-375-3p are elevated in early-onset T1DE1 and that this signal is not solely explained by chronological age or by differences in circulating B cells and CD4^⁺^/CD8^⁺^ T-cell frequencies. Together with prior evidence that miR-150-5p can be induced upon lymphocyte activation and released into the circulation, these findings are consistent with a model in which higher plasma miR-150-5p in T1DE1 reflects qualitative differences in immune states and/or increased miRNA release per cell, rather than simple shifts in lymphocyte abundance, while elevated miR-375-3p may derive from corresponding heightened β-cell death. Consistent with this, we have provided evidence of an additional method for stratification of T1D individuals which relies on measurement of these two circulating miRNAs and can then classify subjects as miRC1 (with a more severe disease) or miRC2. Moving forward, longitudinal validation, mechanistic studies in defined immune subsets, and integrated multi-omic analyses will be required to clarify the causal biology linking miR-150-5p and miR-375-3p to endotype-specific immunopathology and to determine whether and how these markers can be used within precision-medicine approaches for T1D, potentially in combination with other circulating biomarkers (i.e. proinsulin-c peptide ratio)(8).

In conclusion, within emerging frameworks that recognize T1D as a heterogeneous disease, these results underscore the promise of miRNA-based measurements to strengthen patient stratification at diagnosis and may inform enrichment strategies for immunomodulatory trials. Hence, we propose the inclusion of assays for these two circulating miRNAs in future immunotherapy trials.

## Methods

### T1D individuals of the INNODIA first cohort

Circulating small RNA sequencing analysis was performed in two independent cohorts previously described (25) deriving from INNODIA multicentric consortium.

Briefly, the first cohort was composed of 115 individuals with newly diagnosed stage 3 T1D (4.5 ±1.5 weeks from diagnosis) (complete clinical characteristics in **Supplementary Table 1**). The second cohort was composed by n=147 newly diagnosed T1D individuals (3.9±1.8 weeks from diagnosis) (**Supplementary Table 1**). All individuals were positive for at least one autoantibody (GADA, IA-2A, ZnT8A, IAA) and aged between 1-45 years. All individuals (both first and second cohort) were followed-up with programmed visits at 3-(V2), 6-(V3) and 12-months (V4) after diagnosis.

Plasma samples for small RNAs sequencing were collected at baseline (V1, <6 weeks from diagnosis) through a standardised protocol (24) adopted by all clinical sites involved in the multicentric consortium.

### Study approval and Ethics

The study followed the guidelines of the Declaration of Helsinki for research on human individuals, and the study was approved by the local ethical committees of each participating clinical site. Written informed consent was obtained from all participants before their participation in this study.

### Blood samples and plasma processing

Blood samples were collected in K_2_ or K_3_-EDTA 3.5 mL tubes, inverted ten times and stored upright at room temperature (15-25°C), and processed within 2 hours from blood draw. An initial centrifuge was performed at 1800Xg for 10 minutes at 15-25°C to separate blood cells from plasma. Then, plasma was collected in smaller tubes (i.e. 2 mL – Sterile, apyrogen and nuclease-free) avoiding touching the white blood cells interphase (leaving 2-3 mm of plasma layer over the leucocytes) and further centrifuged at 1200Xg for 20 minutes at 10°C to remove contaminant cells and platelets.

Multiple 200 μL plasma aliquots (where possible, 5 aliquots in nuclease-free tubes) were then stored at-80°C and then transferred to a central biobank located in Cambridge (UK) until final transfer to the analytical laboratory.

### QIAseq Small RNA sequencing

Total RNA extraction was performed from 200 µL of plasma through Serum/Plasma Norgen kit (cat. 55000, Thorold, ON L2V 4Y6, Canada). Small RNA-derived cDNA libraries were prepared using QiaSeq miRNA library kit (cat. 331505, Qiagen). QIASeq strategy assign Unique Molecular Index (bound to reverse transcription primers) during reverse transcription step to every mature miRNA molecule, to enable unbiased and accurate miRNome-wide quantification of mature miRNAs by NGS. Then, libraries quality control (QC) was performed quantifying their concentration through QUBIT 3.0 spectrofluorometer (Qubit™ dsDNA HS Assay Kit, cat. Q32854, Thermofisher Scientific) and assessing their quality using capillary electrophoresis in Bioanalyzer 2100 (Agilent High Sensitivity DNA kit cat. 5067-4626, Thermofisher Scientific). High quality of libraries was evaluated considering electropherograms showing a peak comprised between 175 and 185 bp. Following QC, all libraries were normalized until 2 nM and pooled, denatured in 0.2 N NaOH and further sequenced (final concentration 175 pM) on Illumina NovaSeq 6000 platform [NovaSeq 6000 SP Reagent Kit (100 cycles) cat. 20027464, NovaSeq XP 2-Lane Kit cat. 20021664, Illumina] using the XP protocol applying 75×1 single reads.

Data were returned from BaseSpace Sequence Hub as demultiplexed FASTQ files. Resulting raw reads were deduplicated by leveraging Unique Molecular Identifiers (UMIs) present in the library, then mapped to miRbase v21 and piRNABank using QIAseq miRNA Quantification V1 Legacy pipeline from QIAGEN GeneGlobe Data Analysis Center portal (https://geneglobe.qiagen.com/us/analyze). Briefly, resulting reads were mapped referring to miRbase v21 and piRNABank using QIAGEN Gene Globe data analysis center software, which identified a wide repertoire of small RNA species e.g., piRNA (PIWI interacting RNA), tRFs (tRNA fragments), rRNA (ribosomal RNA), miRNA (microRNA).

All these procedures (samples collection strategy and time, RNA extraction, Small RNAs library preparation and sequencing) were also conducted on second cohort as already described for first cohort, with minor modifications. In details, Small RNAs libraries were barcoded with unique dual indexes (UDI) (cat.# 331615 and cat.# 331625).

### MiRNAs differential expression analysis

Reads assigned to miRNAs were standardized into Counts Per Million (CPM) and filtered through *edgeR* package of R (*BioConductor*), maintaining only those miRNAs expressed in at least 70% of individuals with at least 10 CPM. Following low counts filtering, Median of Ratios normalization was performed through *DESeq2* package of R (*BioConductor*) and normalised counts were used for subsequent analyses.

Normalised reads of the two sequencing datasets were used to detect any differentially expressed miRNAs between T1DE1 and T1DE2 groups with *DESeq2* package of R (*BioConductor*) using Wald test and Benjamini-Hoechberg adjusted *P*-value <0.05 was considered as significant.

### Droplet Digital PCR (ddPCR)

Validation of selected miR-150-5p and miR-375-3p, identified through differential expression analysis, was performed through miRCury LNA reverse transcription and subsequent droplet digital PCR (ddPCR) detection.

In details, their expression was analysed in all plasma samples of first and second T1D cohort using miRCury LNA miRNA assay primers (QIAGEN) through a standardised protocol. Briefly, 3µL of RNA (the same used for small RNA sequencing) were reverse transcribed, according to manual’s instruction, by adding 2 µL of 5X miRcury SYBR Green RT Reaction Buffer, 1 µL of 10X miRCury RT Enzyme Mix, 3,5 µL of H_2_O nuclease-free and 0,5 µL UniSp6. The reaction product was incubated at 42 °C for 60 min and then at 95 °C for 5 min. Then, droplet digital PCR was performed on a BioRad QX200 system using a Evagreen assay (BioRad, Mississauga, ON, Canada). Each PCR reaction contained 11 μL of QX200 super mix, 1.1 μL of each miRCury LNA assay primer, 5.9 μL of H2O and 4 μL of template cDNA (diluted 1:10 for miR-375-3p and 1:30 for miR-150-5p evaluation) in a final volume of 22 μL. The PCR reactions were mixed, centrifuged briefly and 20 μL transferred into the sample well of a DG8™ cartridge. After adding 70 μL of QX200™ droplet generation oil into the oil wells, the cartridge was covered using a DG8™ gasket, and droplets were generated using the QX200™ droplet generator. Droplets were carefully transferred into PCR plates using a multi-channel pipette and the plate sealed using PCR plate heat seal foil and the PX1™ PCR plate sealer. PCR was performed in a SimpliAmp thermal cycler (Life technologies, CA, USA). The PCR protocol was 95°C for ‘; 40 cycles of: 95°C for 30”, 56°C 1’ (for both miR-150-5p and miR-375-3p); 4°C for 5’; 90°C for 5’ and hold at 4°C. PCR plates were transferred into a QX200™ droplet reader to count positive and negative droplets. Thresholds to separate positive from negative droplets were set manually for each miRNA using the histogram function and reads analysed using QuantaSoft™ Analysis Pro software (Version 1.2, BioRad, Mississauga, ON, Canada).

### PBMC (cryopreserved) multi-dimensional flow cytometry (Multi-FACS) immunomics

Immunomics profile of peripheral blood from n = 62 out of 115 individuals (only from the INNODIA First Cohort) was investigated at baseline through Cytek Aurora flow cytofluorometer, as previously described (25). Of these 62 individuals, n=10 were included in T1DE1, n=28 in T1DE-I and n=24 in T1DE-I. In order two find differences in the proportion of immune cell populations between the T1DE1 and T1DE2, a Beta regression analysis was performed, using the betareg function (version 3.1.4) The Beta regression model was corrected for the time between blood draw and PBMCs isolation (same day vs. overnight) and then computed as previously described (25).

### Whole-blood Paxgene transcriptomics

Whole-blood RNA-sequencing data were available for n= 49/115 INNODIA T1D individuals belonging to the First Cohort. RNA-seq was performed as previously described (77). Briefly, frozen whole-blood PAXgene samples were thawed at room temperature for 2 h and subjected to RNA extraction using PAXgene Blood miRNA Kit. Total RNA was purified, following the protocol supplied by the kit manufacturer. RNA-seq libraries were prepared using TruSeq stranded mRNA HT kit and protocol. Pooled libraries were sequenced on an Illumina NovaSeq 6000 instrument, using 2×50 bp paired-end sequencing. Overexpressed haemoglobin-related genes were filtered prior to normalisation. The data were CPM scaled with TMM normalisation factors using the R package edgeR, then log_2_ transformed. Only genes with an average CPM > 1 across all samples were included in the analysis.

### Tissue expression analysis

To evaluate the human tissue and cell-type expression profile of miR-150-5p, we performed an in-silico analysis using isomiRdb (https://ccb-compute.cs.uni-saarland.de/isomirdb), a publicly available compendium of uniformly processed human small RNA–sequencing datasets. isomiRdb aggregates miRNA/isomiR expression values from 42,499 miRNA-seq samples (including datasets from miRMaster, TCGA, and SRA) processed through a standardized pipeline (miRMaster/sRNAbench) with miRNA annotation based on miRBase v22.1. miR-150-5p expression was queried at the miRNA level and summarized across the database’s curated metadata for tissues and cell types, extracting the reported normalized expression values (RPM) for comparative visualization and qualitative assessment of tissue and cellular enrichment.

## Statistical Analyses

Statistical analyses were performed in R (v4.2.2) or Python 3.9, and GraphPad Prism (v10.0). Exact P values or P values thresholds, and the number of observations contributing to each analysis are reported in the corresponding figures/tables legends.

For circulating miRNA discovery, small RNA-seq read counts were first converted to counts per million (CPM) and low-abundance features were removed by retaining only miRNAs detected in ≥70% of individuals with ≥10 CPM, after which median-of-ratios normalization was applied (DESeq2). Differential expression between T1DE1 vs T1DE2 was assessed within each cohort using DESeq2 (Wald test), controlling for multiple testing with the Benjamini–Hochberg (BH) false discovery rate; an adjusted P<0.05 was considered significant for the primary miRNA differential-expression screen. ddPCR quantification for miR-150-5p and miR-375-5p (reported as copies/µL on log scale in the figures) and other non-normally distributed outcomes were compared across endotypes (T1DE1, T1DEInt, T1DE2) using Kruskal–Wallis tests followed by multiple-comparison procedures (Dunn’s test) as specified in the relevant figure legends.

Cross-sectional data at baseline-Visit 1, for comparisons across the three age-defined endotypes (T1DE1, T1DEInt, T1DE2) or between miR-combined subgroups (miRC1 vs miRC2) were evaluated using the Kruskal–Wallis test (Table 1 and Table 2) or two-sided Mann–Whitney U tests (for continuous variables) (Table 5). When the global Kruskal–Wallis test was statistically significant, post hoc pairwise comparisons were performed using Dunn’s multiple comparison test. Categorical variables, including sex distribution and autoantibody positivity rates (defined according to pre-specified assay-specific cutoffs), were compared using the chi-square test for overall comparisons across three groups and two-sided Fisher’s exact test for pairwise (Table 1 and Table 2) or two-group analyses (Table 5). To account for multiple hypothesis testing, false discovery rate (FDR) correction was applied using the Benjamini–Hochberg procedure. Exact P values <0.1 were shown in Tables 1-2 and 5, otherwise *n.s*. were indicated.

Longitudinal metabolic outcomes collected at V1 (<6 weeks from diagnosis), V2 (3 months), V3 (6 months) and V4 (12 months) were analysed using a mixed-effects model for repeated measures (to accommodate unbalanced follow-up), applying Geisser–Greenhouse correction when sphericity could not be assumed and Tukey’s-adjusted post hoc testing for pairwise endotype contrasts at specific visits. We relied on the repeated measures mixed-effects model to incorporate missing observations under a missing at random assumption (we had no evidence that missing observation were systematically different from those available). Fixed-effects were the endotype (T1DE1, T1DE2, T1DEInt), visits (V1-V4, and the endotype × Visit interaction. Random-effects (to account for repeated measures) were the subject (random intercept) and the residual error term.

C-peptide decline over the first year was quantified from stimulated β-cell function as the percentage change in Mixed Meal Tolerance Test (MTTT) AUC C-peptide from V2 to V4, computed as %Δ = ((AUC_V2 − AUC_V4)/AUC_V2) × 100, and compared across T1DE endotypes using Kruskal–Wallis plus Dunn’s multiple comparison tests; values are summarized as median and Interquartile Range (IQR). MMTT test was not performed at V1 in the INNODIA study.

Correlation analyses were performed using Spearman’s rho; for integrative analyses with whole-blood transcriptomics, miR-150-5p (ddPCR copies/uL) was correlated with gene-expression values (log2-transformed normalized reads), and transcript-level multiple testing was controlled by FDR-adjusted P values. Where indicated, normalized expression values were further transformed (log2 or Z-score) for visualization and comparability across analyses. Publicly available raw data on cohorts deriving from non-diabetic, coeliac disease and childhood asthma individuals were used as control datasets to investigate the specificity of differentially expressed miRNAs in T1D age-endotypes and were re-analysed starting from raw FASTQ files, where available, and analyused as reported above.

Discriminatory performance of single-miRNA (miR-150-5p or miR-375-3p ddPCR copies/uL) measurements for endotype classification (T1DE1 – T1DE2) was evaluated by receiver operating characteristic (ROC) analysis (AUC with 95% CI) and performed separately in each cohort or on the totality of the population (first + second cohort). Statistical significance of AUCs was assessed against the null expectation (AUC = 0.5) using GraphPad Prism v.10. Sensitivity and specificity values were selected by Youden index (J) calculation [ J= (Sensitivity + Specificity)-1] to identify the best discriminatory cutoff (corresponding to J_max_) for each miRNA.

Receiver operating characteristic (ROC) curves for combined miRNAs were constructed using a combined miRNA score obtained from a logistic regression model integrating miR-150-5p+miR-375-3p expression levels, with T1DE1 or T1DE2 used as the outcome. ROC curves were analysed as reported above.

To re-classify T1D individuals using both miRNAs in an interpretable model, we used the best T1DE1 vs T1DE2 discriminatory cutoffs for each miRNA, as calculated on the totality of the T1D population (T1DE1 and T1DE2 individuals of the first cohort + second cohort). Hence, based on these cutoffs, a combined miRNAs signature was defined using a logical “AND” rule, whereby subjects were classified as miRNA-high (miRC1) only if both miR-150-5p and miR-375-3p satisfied their respective Youden index-derived cutoffs (≥18.95 and ≥6.7 copies/µL of plasma, respectively) (**Supplemental Figure 4A and 4B**); all remaining subjects were classified as miRNA-low (miRC2) (**Figure 5C**). This rule was subsequently applied to the full study population (first + second cohort), including subjects belonging to the intermediate age endotype T1DE-Int, to enable miRNA-based re-stratification across disease stages.

## Data and code availability

Circulating Small RNA-seq data from T1D individuals of the first and second cohort are available in Gene Expression Omnibus (GEO): GSE265980 (First cohort), GSE265981 (Second cohort) (25). Circulating Small RNA-seq data from subjects with childhood asthma are available in Gene Expression Omnibus (GEO): GSE134897 (39), while data of small RNA-seq from individuals with Coeliac disease are available as NCBI BioProject repository with the identification code PRJNA793584 (38).

Any additional information required to reanalyze the data reported in this work is available from the lead contact upon request.

## Author Contributions

G.E.G. and G.S. contributed to the experimental design, performed experimental work and data analysis and interpretation, and drafted the manuscript.

E.P. and G.L. contributed to droplet digital PCR validation and data acquisition, and data interpretation.

T.S., I.S., L.E. and R.L. performed experimental work, bioinformatic analyses and data interpretation for the integration of transcriptomic datasets.

T.T. performed experimental work, bioinformatic analyses and data interpretation for the integration of immunomic datasets.

P.L., S.J.R. and N.G.M. originally defined the age-related T1D endotypes and provided the conceptual framework that inspired the present miRNA-based endotype analysis. They contributed to data interpretation and critically revised the manuscript

F.D. and G.S. conceived and supervised the study, secured funding, contributed to data interpretation, and critically revised the manuscript.

All authors reviewed and approved the final version of the manuscript.

## Data Availability

Circulating Small RNA-seq data from T1D individuals of the first and second cohort are available in Gene Expression Omnibus (GEO): GSE265980 (First cohort), GSE265981 (Second cohort) (25). Circulating Small RNA-seq data from subjects with childhood asthma are available in Gene Expression Omnibus (GEO): GSE134897 (39), while data of small RNA-seq from individuals with Coeliac disease are available as NCBI BioProject repository with the identification code PRJNA793584 (38).
Any additional information required to reanalyze the data reported in this work is available from the lead contact upon request.

## Acknowledgements

The secretarial help of Alessandra Mechini was highly appreciated.

## References

1. American Diabetes Association Professional Practice Committee for Diabetes*. 2. Diagnosis and Classification of Diabetes: Standards of Care in Diabetes-2026. Diabetes Care. 2026;49(Supplement_1):S27–S49.

2. Redondo MJ, Morgan NG. Heterogeneity and endotypes in type 1 diabetes mellitus. Nat Rev Endocrinol. 2023;19(9):542–554.

3. Nigi L, et al. From immunohistological to anatomical alterations of human pancreas in type 1 diabetes: New concepts on the stage. Diabetes Metab Res Rev. 2020;36(4):e3264.

4. Leete P, et al. The effect of age on the progression and severity of type 1 diabetes: potential effects on disease mechanisms. Curr Diab Rep. 2018;18(11):115.

5. Weston CS, et al. Type 1 diabetes: A new vision of the disease based on endotypes. Diabetes Metab Res Rev. 2024;40(2):e3770.

6. Battaglia M, et al. Introducing the endotype concept to address the challenge of disease heterogeneity in type 1 diabetes. Diabetes Care. 2020;43(1):5–12.

7. Leete P, et al. Differential Insulitic Profiles Determine the Extent of β-Cell Destruction and the Age at Onset of Type 1 Diabetes. Diabetes. 2016;65(5):1362–1369.

8. Leete P, et al. Studies of insulin and proinsulin in pancreas and serum support the existence of aetiopathological endotypes of type 1 diabetes associated with age at diagnosis. Diabetologia. 2020;63(6):1258–1267.

9. Arif S, et al. Blood and islet phenotypes indicate immunological heterogeneity in type 1 diabetes. Diabetes. 2014;63(11):3835–3845.

10. Torabi F, et al. Differential expression of genes controlling lymphocyte differentiation and migration in two distinct endotypes of type 1 diabetes. Diabet Med. 2023;40(9):e15155.

11. Carr ALJ, et al. Circulating C-Peptide Levels in Living Children and Young People and Pancreatic β-Cell Loss in Pancreas Donors Across Type 1 Diabetes Disease Duration. Diabetes. 2022;71(7):1591–1596.

12. Fiorina P, Pozzilli P. Unveiling a novel type 1 diabetes endotype: Opportunities for intervention. Diabetes Metab Res Rev. 2022;38(5):e3536.

13. Wilson MAR, Pozzilli P. What type 1 diabetes endotype is most suitable for anti-CD3 antibodies prevention trials? J Diabetes Complicat. 2025;39(10):109132.

14. Sebastiani G, et al. Circulating microRNAs and diabetes mellitus: a novel tool for disease prediction, diagnosis, and staging? J Endocrinol Invest. 2017;40(6):591–610.

15. Dotta F, et al. MicroRNAs: markers of β-cell stress and autoimmunity. Curr Opin Endocrinol Diabetes Obes. 2018;25(4):237–245.

16. O’Connell RM, et al. Physiological and pathological roles for microRNAs in the immune system. Nat Rev Immunol. 2010;10(2):111–122.

17. Grieco GE, et al. The Landscape of microRNAs in βCell: Between Phenotype Maintenance and Protection. Int J Mol Sci. 2021;22(2). 10.3390/ijms22020803.

18. Guay C, et al. Lymphocyte-Derived Exosomal MicroRNAs Promote Pancreatic β Cell Death and May Contribute to Type 1 Diabetes Development. Cell Metab. 2019;29(2):348–361.e6.

19. Aljani B, et al. Small RNA-Seq and real time rt-qPCR reveal islet miRNA released under stress conditions. Islets. 2024;16(1):2392343.

20. Lakhter AJ, et al. Beta cell extracellular vesicle miR-21-5p cargo is increased in response to inflammatory cytokines and serves as a biomarker of type 1 diabetes. Diabetologia. 2018;61(5):1124–1134.

21. Dekkers MC, et al. Extracellular vesicles derived from stressed beta cells mediate monocyte activation and contribute to islet inflammation. Front Immunol. 2024;15:1393248.

22. Guay C, et al. Horizontal transfer of exosomal microRNAs transduce apoptotic signals between pancreatic beta-cells. Cell Commun Signal. 2015;13:17.

23. Suomi T, et al. Type 1 Diabetes in Children With Genetic Risk May Be Predicted Very Early With a Blood miRNA. Diabetes Care. 2022;45(4):e77–e79.

24. Grieco GE, et al. Protocol to analyze circulating small non-coding RNAs by high-throughput RNA sequencing from human plasma samples. STAR Protocols. 2021;2(3):100606.

25. Sebastiani G, et al. A set of circulating microRNAs belonging to the 14q32 chromosome locus identifies two subgroups of individuals with recent-onset type 1 diabetes. Cell Rep Med. 2024;5(6):101591.

26. Ventriglia G, et al. miR-409-3p is reduced in plasma and islet immune infiltrates of NOD diabetic mice and is differentially expressed in people with type 1 diabetes. Diabetologia. 2020;63(1):124–136.

27. Sassi G, et al. A Plasma miR-193b-365 Signature Combined With Age and Glycemic Status Predicts Response to Lactococcus lactis-Based Antigen-Specific Immunotherapy in New-Onset Type 1 Diabetes. Diabetes. 2023;72(10):1470–1482.

28. Formichi C, et al. Circulating microRNAs Signature for Predicting Response to GLP1-RA Therapy in Type 2 Diabetic Patients: A Pilot Study. Int J Mol Sci. 2021;22(17). 10.3390/ijms22179454.

29. Marcovecchio ML, et al. The INNODIA Type 1 Diabetes Natural History Study: a European cohort of newly diagnosed children, adolescents and adults. Diabetologia. 2024;67(6):995–1008.

30. Dunger DB, et al. INNODIA Master Protocol for the evaluation of investigational medicinal products in children, adolescents and adults with newly diagnosed type 1 diabetes. Trials. 2022;23(1):414.

31. Gomez-Muñoz L, et al. Identification of peripheral blood endotypes associated with age in pediatric type 1 diabetes. Pediatr Diabetes. 2025;2025:5512196.

32. Parviainen A, et al. Heterogeneity of Type 1 Diabetes at Diagnosis Supports Existence of Age-Related Endotypes. Diabetes Care. 2022;45(4):871–879.

33. Zhou Q, et al. Phenotypic Spectrum at Diagnosis of Age-Related Endotypes of Type 1 Diabetes Mellitus: A Cross-Sectional Study in China. J Diabetes. 2025;17(6):e70111.

34. Sales Luis M, et al. Children with type 1 diabetes of early age at onset - immune and metabolic phenotypes. J Pediatr Endocrinol Metab. 2019;32(9):935–941.

35. Achenbach P, et al. A classification and regression tree analysis identifies subgroups of childhood type 1 diabetes. EBioMedicine. 2022;82:104118.

36. Özer E, et al. Pancreatic Ultrasound Features at Diagnosis of Type 1 Diabetes: Age-Related Differences in Children. J Clin Med. 2025;14(21). 10.3390/jcm14217490.

37. Gao S-Y, et al. Age-related heterogeneity of type 1 diabetes mellitus in children: a single-center retrospective study. World J Pediatr. [published online ahead of print: December 27, 2025]. 10.1007/s12519-025-01004-3.

38. Felli C, et al. Circulating microRNAs as novel non-invasive biomarkers of paediatric celiac disease and adherence to gluten-free diet. EBioMedicine. 2022;76:103851.

39. Li J, et al. Circulating micrornas and treatment response in childhood asthma. Am J Respir Crit Care Med. 2020;202(1):65–72.

40. Aparicio-Puerta E, et al. isomiRdb: microRNA expression at isoform resolution. Nucleic Acids Res. 2023;51(D1):D179–D185.

41. Joglekar MV, et al. Expression of islet-specific microRNAs during human pancreatic development. Gene Expr Patterns. 2009;9(2):109–113.

42. Avnit-Sagi T, et al. The promoter of the pri-miR-375 gene directs expression selectively to the endocrine pancreas. PLoS ONE. 2009;4(4):e5033.

43. Poy MN, et al. A pancreatic islet-specific microRNA regulates insulin secretion. Nature. 2004;432(7014):226–230.

44. Kredo-Russo S, et al. Pancreas-enriched miRNA refines endocrine cell differentiation. Development. 2012;139(16):3021–3031.

45. Zhou B, et al. miR-150, a microRNA expressed in mature B and T cells, blocks early B cell development when expressed prematurely. Proc Natl Acad Sci USA. 2007;104(17):7080–7085.

46. Ying W, et al. miR-150 regulates obesity-associated insulin resistance by controlling B cell functions. Sci Rep. 2016;6:20176.

47. Xiao C, et al. MiR-150 controls B cell differentiation by targeting the transcription factor c-Myb. Cell. 2007;131(1):146–159.

48. Ménoret A, et al. Antigen-specific downregulation of miR-150 in CD4 T cells promotes cell survival. Front Immunol. 2023;14:1102403.

49. Ban YH, et al. miR-150-Mediated Foxo1 Regulation Programs CD8+ T Cell Differentiation. Cell Rep. 2017;20(11):2598–2611.

50. Chen Z, et al. miR-150 Regulates Memory CD8 T Cell Differentiation via c-Myb. Cell Rep. 2017;20(11):2584–2597.

51. Evans-Molina C, et al. The heterogeneity of type 1 diabetes: implications for pathogenesis, prevention, and treatment-2024 Diabetes, Diabetes Care, and Diabetologia Expert Forum. Diabetologia. 2025;68(9):1859–1878.

52. Morgan NG. Insulitis in human type 1 diabetes: lessons from an enigmatic lesion. Eur J Endocrinol. 2024;190(1):R1–9.

53. Starskaia I, et al. Distinct cellular immune responses in children en route to type 1 diabetes with different first-appearing autoantibodies. Nat Commun. 2024;15(1):3810.

54. Guay C, Regazzi R. Circulating microRNAs as novel biomarkers for diabetes mellitus. Nat Rev Endocrinol. 2013;9(9):513–521.

55. Erener S, et al. Profiling of circulating microRNAs in children with recent onset of type 1 diabetes. JCI Insight. 2017;2(4):e89656.

56. Bravo-Egana V, et al. Quantitative differential expression analysis reveals miR-7 as major islet microRNA. Biochem Biophys Res Commun. 2008;366(4):922–926.

57. Poy MN, et al. miR-375 maintains normal pancreatic alpha-and beta-cell mass. Proc Natl Acad Sci USA. 2009;106(14):5813–5818.

58. Sebastiani G, et al. MicroRNA expression profiles of human iPSCs differentiation into insulin-producing cells. Acta Diabetol. 2017;54(3):265–281.

59. Grieco GE, et al. Leveraging circulating microRNAs for personalized disease-modifying therapies in type 1 diabetes. Per Med. 2025;1–13.

60. Latreille M, et al. miR-375 gene dosage in pancreatic β-cells: implications for regulation of β-cell mass and biomarker development. J Mol Med. 2015;93(10):1159–1169.

61. Martens GA, et al. The microrna landscape of acute beta cell destruction in type 1 diabetic recipients of intraportal islet grafts. Cells. 2021;10(7). 10.3390/cells10071693.

62. Piemonti L, et al. Induction of immune education in type 1 diabetes through controlled allogeneic islet rejection at onset: a monocentric open-label pilot study. EClinicalMedicine. 2025;90:103685.

63. Song I, et al. Circulating microRNA-375 as biomarker of pancreatic beta cell death and protection of beta cell mass by cytoprotective compounds. PLoS ONE. 2017;12(10):e0186480.

64. Roat R, et al. Identification and Characterization of microRNAs Associated With Human β-Cell Loss in a Mouse Model. Am J Transplant. 2017;17(4):992–1007.

65. Erener S, et al. Circulating miR-375 as a biomarker of β-cell death and diabetes in mice. Endocrinology. 2013;154(2):603–608.

66. Roat R, et al. Circulating miRNA-150-5p is associated with immune-mediated early β-cell loss in a humanized mouse model. Xenotransplantation. 2019;26(2):e12474.

67. Leete P. Type 1 diabetes in the pancreas: A histological perspective. Diabet Med. 2023;40(12):e15228.

68. Monticelli S, et al. MicroRNA profiling of the murine hematopoietic system. Genome Biol. 2005;6(8):R71.

69. Allantaz F, et al. Expression profiling of human immune cell subsets identifies miRNA-mRNA regulatory relationships correlated with cell type specific expression. PLoS ONE. 2012;7(1):e29979.

70. Hu Y-Z, et al. Multiple functions and regulatory network of miR-150 in B lymphocyte-related diseases. Front Oncol. 2023;13:1140813.

71. de Candia P, et al. Intracellular modulation, extracellular disposal and serum increase of MiR-150 mark lymphocyte activation. PLoS ONE. 2013;8(9):e75348.

72. de Candia P, et al. Serum microRNAs as Biomarkers of Human Lymphocyte Activation in Health and Disease. Front Immunol. 2014;5:43.

73. Torri A, et al. Extracellular microrna signature of human helper T cell subsets in health and autoimmunity. J Biol Chem. 2017;292(7):2903–2915.

74. Xiao C, et al. MiR-150 Controls B Cell Differentiation by Targeting the Transcription Factor c-Myb. Cell. 2016;165(4):1027.

75. Ghisi M, et al. Modulation of microRNA expression in human T-cell development: targeting of NOTCH3 by miR-150. Blood. 2011;117(26):7053–7062.

76. Smith NL, et al. miR-150 Regulates Differentiation and Cytolytic Effector Function in CD8+ T cells. Sci Rep. 2015;5:16399.

77. Suomi T, et al. Gene expression signature predicts rate of type 1 diabetes progression. EBioMedicine. 2023;92:104625.

