## Supplementary material for "Circulating plasma microRNAs miR-150 and miR-375 levels are associated with age-related endotypes of newly diagnosed Type 1 Diabetes"

**SUPPLEMENTAL MATERIAL**

### Supplemental Tables

**Supplemental Table 1.** Demographics and clinical characteristics of T1DM individuals belonging to the First and Second INNODIA cohort. Numbers of T1DM people with available clinical parameter are reported in brackets. Values are reported as mean ± S.D for continuous variables.

| Demographics/ Clinical characteristics | First T1DM cohort | Second T1DM cohort |
|---|---|---|
| Age (years) | 12.51 ± 7.70 [115] | 12.03 ± 7.82 [147] |
| Age range (year) | 2-38 [115] | 1-42 [147] |
| Sex (Female/Male) | 58F/57M | 55F/92M |
| BMI (Kg/$m^2$) | 23.12 ± 2.82 [16] | 22.26 ± 2.99 [21] |
| BMI_SDS | 0.15 ± 1.09 [99] | 0.38 ± 1.12 [126] |
| Disease duration (weeks) | 4.08 ± 1.53 [115] | 3.91 ± 1.76 [142] |
| Fasting C-peptide (pmol/L) | 277.64 ± 203.54 [115] | 270.03 ± 194.79 [145] |
| HbA1c (mmol/mol) | 77.45 ± 19.44 [112] | 76.01 ± 19.26 [143] |
| Insulin dose (units/kg/day) | 0.52 ± 0.27 [113] | 0.59 ± 0.40 [144] |
| IAA (% positive) | 76 [88] | 77.55 [114] |
| IA2A (% positive) | 72,1 [83] | 78.23 [115] |
| GADA (% positive) | 76 [88] | 77.55 [114] |
| ZnT8A (% positive) | 66 [76] | 70.75 [104] |

**Supplemental Table 2A.** Differentially Expressed Circulating miRNAs between T1DE1 (n=18) and T1DE2 (n=38) endotypes individuals in the first INNODIA cohort. miRNA are ordered based on their significance (from the smallest to the largest, Padj < 0.05). Highlighted in green: miR-150-5p and miR-375-3p which are shared (both in terms of significance and direction of the Fold Change) between first and second INNODIA cohort.

| miRNA | Normalized Reads Counts | log2 Fold Change T1DE1 vs T1DE2 | lfcSE | stat | pvalue | padj (FDR) |
|---|---|---|---|---|---|---|
| miR-150-5p | 2793,88 | 0,87 | 0,15 | 5,85 | 4,91E-09 | 1,17E-06 |
| miR-140-5p | 472,57 | 1,29 | 0,25 | 5,11 | 3,16E-07 | 3,76E-05 |
| miR-181a-3p | 48,65 | 1,28 | 0,28 | 4,53 | 5,88E-06 | 4,67E-04 |
| miR-181b-5p | 380,42 | 0,83 | 0,21 | 4,04 | 5,27E-05 | 3,14E-03 |
| miR-155-5p | 734,06 | 1,27 | 0,33 | 3,84 | 1,22E-04 | 5,82E-03 |
| miR-320d | 64,83 | 0,50 | 0,14 | 3,64 | 2,77E-04 | 1,10E-02 |
| let-7f-5p | 57335,73 | 1,40 | 0,41 | 3,40 | 6,73E-04 | 1,98E-02 |
| miR-181a-5p | 2007,56 | 0,80 | 0,24 | 3,34 | 8,32E-04 | 1,98E-02 |
| miR-215-5p | 144,49 | 0,74 | 0,22 | 3,42 | 6,17E-04 | 1,98E-02 |
| miR-23b-3p | 853,58 | 1,04 | 0,31 | 3,36 | 7,87E-04 | 1,98E-02 |
| miR-224-5p | 574,18 | 1,89 | 0,57 | 3,32 | 9,16E-04 | 1,98E-02 |
| miR-21-5p | 17284,44 | 0,88 | 0,27 | 3,27 | 1,06E-03 | 2,02E-02 |
| miR-941 | 92,98 | 0,84 | 0,26 | 3,26 | 1,10E-03 | 2,02E-02 |
| miR-375 | 263,66 | 0,74 | 0,23 | 3,14 | 1,66E-03 | 2,82E-02 |
| miR-199b-5p | 24,41 | 0,62 | 0,20 | 3,11 | 1,88E-03 | 2,98E-02 |
| miR-125b-5p | 671,75 | 0,51 | 0,17 | 3,01 | 2,58E-03 | 3,61E-02 |
| miR-29a-3p | 1160,76 | 0,46 | 0,15 | 3,03 | 2,45E-03 | 3,61E-02 |
| miR-10a-5p | 725,10 | 0,59 | 0,20 | 2,96 | 3,03E-03 | 3,91E-02 |
| miR-181a-2-3p | 87,41 | 0,93 | 0,32 | 2,95 | 3,13E-03 | 3,91E-02 |
| miR-4446-3p | 35,23 | 1,05 | 0,36 | 2,94 | 3,28E-03 | 3,91E-02 |
| miR-103a-3p | 11638,03 | 0,98 | 0,34 | 2,91 | 3,63E-03 | 4,12E-02 |
| miR-222-3p | 928,61 | 1,08 | 0,37 | 2,89 | 3,90E-03 | 4,22E-02 |
| miR-664a-3p | 61,97 | 0,53 | 0,19 | 2,84 | 4,54E-03 | 4,70E-02 |
| miR-130b-5p | 56,20 | 1,02 | 0,37 | 2,79 | 5,20E-03 | 4,93E-02 |
| miR-146b-5p | 719,50 | 1,13 | 0,41 | 2,77 | 5,66E-03 | 4,93E-02 |
| miR-193a-5p | 134,70 | 0,44 | 0,16 | 2,75 | 6,01E-03 | 4,93E-02 |
| miR-320b | 364,90 | 0,45 | 0,16 | 2,76 | 5,82E-03 | 4,93E-02 |
| miR-320c | 218,42 | 0,47 | 0,17 | 2,78 | 5,39E-03 | 4,93E-02 |
| miR-505-3p | 32,37 | 0,76 | 0,27 | 2,79 | 5,27E-03 | 4,93E-02 |

**Supplemental Table 2B.** Differentially Expressed Circulating miRNAs (Padj<0.05) between T1DE1 (n=30) and T1DE2 (n=57) endotypes individuals in the second INNODIA cohort. miRNAs are ordered based on their significance (from the smallest to the largest, Padj < 0.05). Highlighted in green: miR-150-5p and miR-375-3p which are shared (both in terms of significance and direction of the Fold Change) between first and second INNODIA cohort.

| miRNA | Normalized Reads Counts | log2FoldChange T1DE1 vs T1DE2 | lfcSE | stat | pvalue | padj (FDR) |
|---|---|---|---|---|---|---|
| miR-150-5p | 596,44 | 1,30 | 0,21 | 6,18 | 6,49E-10 | 1,47E-07 |
| miR-335-5p | 2478,13 | -1,63 | 0,32 | -5,05 | 4,52E-07 | 5,10E-05 |
| miR-340-5p | 383,40 | -1,52 | 0,32 | -4,75 | 2,06E-06 | 1,55E-04 |
| miR-99b-5p | 331,82 | -1,43 | 0,31 | -4,56 | 5,08E-06 | 2,87E-04 |
| miR-221-3p | 960,67 | -1,57 | 0,35 | -4,45 | 8,54E-06 | 3,86E-04 |
| let-7e-5p | 2248,16 | -1,42 | 0,33 | -4,33 | 1,52E-05 | 5,58E-04 |
| miR-146a-5p | 424,71 | -1,50 | 0,35 | -4,24 | 2,22E-05 | 5,58E-04 |
| miR-151a-3p | 2085,96 | -1,47 | 0,34 | -4,27 | 1,94E-05 | 5,58E-04 |
| miR-744-5p | 2288,06 | -1,59 | 0,37 | -4,26 | 2,07E-05 | 5,58E-04 |
| miR-126-5p | 19539,33 | -1,15 | 0,28 | -4,05 | 5,17E-05 | 1,17E-03 |
| miR-374b-5p | 105,61 | -1,39 | 0,35 | -3,97 | 7,28E-05 | 1,50E-03 |
| miR-191-3p | 92,31 | -0,84 | 0,22 | -3,85 | 1,16E-04 | 2,18E-03 |
| miR-181c-3p | 83,42 | -0,81 | 0,22 | -3,68 | 2,37E-04 | 4,02E-03 |
| miR-2392 | 173,71 | -0,74 | 0,20 | -3,64 | 2,71E-04 | 4,02E-03 |
| miR-338-3p | 60,59 | -0,79 | 0,22 | -3,64 | 2,73E-04 | 4,02E-03 |
| miR-4446-3p | 54,95 | -1,07 | 0,30 | -3,63 | 2,84E-04 | 4,02E-03 |
| miR-1301-3p | 386,41 | -1,06 | 0,30 | -3,57 | 3,54E-04 | 4,21E-03 |
| miR-24-3p | 5134,70 | -1,18 | 0,33 | -3,59 | 3,36E-04 | 4,21E-03 |
| miR-4433b-5p | 202,75 | -1,27 | 0,35 | -3,60 | 3,17E-04 | 4,21E-03 |
| miR-199a-3p | 2091,91 | -1,24 | 0,35 | -3,54 | 3,94E-04 | 4,46E-03 |
| let-7f-5p | 79978,01 | -1,05 | 0,30 | -3,49 | 4,77E-04 | 4,98E-03 |
| miR-126-3p | 83126,15 | -1,06 | 0,30 | -3,48 | 4,93E-04 | 4,98E-03 |
| miR-766-3p | 68,69 | -0,67 | 0,19 | -3,48 | 5,07E-04 | 4,98E-03 |
| miR-26a-5p | 65082,63 | -1,21 | 0,35 | -3,45 | 5,61E-04 | 5,29E-03 |
| miR-27b-3p | 162,16 | -0,77 | 0,22 | -3,42 | 6,35E-04 | 5,74E-03 |
| miR-409-3p | 337,03 | -1,32 | 0,39 | -3,41 | 6,61E-04 | 5,74E-03 |
| miR-501-3p | 53,03 | 0,54 | 0,16 | 3,38 | 7,35E-04 | 6,16E-03 |
| miR-125a-5p | 267,66 | -0,80 | 0,24 | -3,35 | 8,09E-04 | 6,50E-03 |
| miR-375 | 853,20 | 0,61 | 0,18 | 3,34 | 8,34E-04 | 6,50E-03 |
| miR-27a-3p | 419,16 | -0,87 | 0,26 | -3,27 | 1,07E-03 | 7,77E-03 |
| miR-625-5p | 606,24 | -1,17 | 0,36 | -3,28 | 1,04E-03 | 7,77E-03 |
| let-7d-5p | 28359,74 | -0,95 | 0,29 | -3,23 | 1,22E-03 | 8,64E-03 |
| miR-139-3p | 220,19 | -0,83 | 0,26 | -3,16 | 1,59E-03 | 1,09E-02 |
| let-7i-5p | 111273,93 | -0,82 | 0,27 | -3,04 | 2,40E-03 | 1,51E-02 |
| miR-1306-5p | 87,51 | -0,42 | 0,14 | -3,05 | 2,29E-03 | 1,51E-02 |
| miR-30d-5p | 14999,15 | -0,93 | 0,31 | -3,04 | 2,40E-03 | 1,51E-02 |
| miR-223-3p | 3296,14 | -1,05 | 0,35 | -2,96 | 3,03E-03 | 1,85E-02 |
| miR-18a-5p | 438,63 | -0,75 | 0,27 | -2,84 | 4,53E-03 | 2,69E-02 |
| miR-766-5p | 48,77 | -0,62 | 0,22 | -2,79 | 5,21E-03 | 3,02E-02 |
| miR-181d-5p | 63,03 | -0,70 | 0,25 | -2,76 | 5,82E-03 | 3,29E-02 |

| | | | | | | |
|---|---|---|---|---|---|---|
| miR-223-5p | 686,05 | -0,81 | 0,30 | -2,73 | 6,28E-03 | 3,46E-02 |
| miR-23a-3p | 723,33 | -0,74 | 0,27 | -2,69 | 7,09E-03 | 3,81E-02 |
| miR-143-3p | 6538,76 | -0,72 | 0,27 | -2,68 | 7,34E-03 | 3,84E-02 |
| miR-328-3p | 63,23 | -0,38 | 0,14 | -2,67 | 7,48E-03 | 3,84E-02 |
| miR-382-5p | 928,48 | -0,92 | 0,35 | -2,65 | 7,97E-03 | 4,00E-02 |
| miR-361-5p | 886,89 | -0,66 | 0,25 | -2,61 | 9,06E-03 | 4,39E-02 |
| miR-652-3p | 2119,33 | -0,88 | 0,34 | -2,61 | 9,13E-03 | 4,39E-02 |

**Supplemental Table 3.** Linear regression estimate and p-values of miR-150-5p and miR-375-3p vs age at onset in multiple cohorts analysed in the study (i.e. INNODIA T1DM first and second cohort, healthy controls, Asthma, Celiac disease).

| miRNA | cohort | Number | Methodology | Rho | Pvalue |
|---|---|---|---|---|---|
| miR-150-5p | First T1DM | 115 | seq | -0,41 | 0,0005 |
| miR-150-5p | Second- T1DM | 147 | seq | -0,36 | 0,0005 |
| miR-150-5p | First T1DM | 109 | ddPCR | -0,25 | 0,037 |
| miR-150-5p | Second T1DM | 147 | ddPCR | -0,17 | 0,0071 |
| miR-375-3p | First T1DM | 115 | seq | -0,37 | <0,0001 |
| miR-375-3p | Second- T1DM | 147 | seq | -0,28 | 0,0005 |
| miR-375-3p | First T1DM | 109 | ddPCR | -0,29 | 0,016 |
| miR-375-3p | Second T1DM | 147 | ddPCR | -0,17 | 0,05 |
| miR-150-5p | Non diabetic/healthy controls | 28 | seq | 0,27 | 0,15 |
| miR-375-3p | Non diabetic/healthy controls | 28 | seq | -0,12 | 0,52 |
| miR-150-5p | Children Asthma | 274 | seq | -0,04 | 0,5 |
| miR-375-3p | Children Asthma | 274 | seq | -0,04 | 0,43 |
| miR-150-5p | Celiac Disease | 23 | seq | -0,12 | 0,58 |
| miR-375-3p | Celiac Disease | 23 | seq | -0,23 | 0,26 |

**Supplemental Table 4.** Details of TargetScan 7.2 analysis of miR-150-5p target genes MPPE1 and RABGAP1L.

| | Predicted consequential pairing of target region (top) and miRNA (bottom) | Site type | Context++ score | Context++ score percentile | Weighted context++ score | Conserved branch length | $P_{CT}$ | Predicted relative $K_D$ |
|---|---|---|---|---|---|---|---|---|
| Position 493-499 of MPPE1 3' UTR | 5' ...UGUAAUCCCAGCACUUUGGGAGG... | | | | | | | |
| | \|\|\|\|\|\|\| | 7mer-m8 | -0.02 | 37 | -0.01 | 0.215 | < 0.1 | -0.943 |
| hsa-miR-150-5p | 3' GUGACCAUGUUCCCAACCCUCU | | | | | | | |
| Position 2194-2200 of RABGAP1L 3' UTR | 5' ...UGUAAUCCCAGCACUUUGGGAGG... | | | | | | | |
| | \|\|\|\|\|\|\| | 7mer-m8 | -0.02 | 37 | -0.02 | 0.154 | < 0.1 | -0.943 |
| hsa-miR-150-5p | 3' GUGACCAUGUUCCCAACCCUCU | | | | | | | |

**Supplemental Figures**

**Supplemental Figure 1.** miR-150-5p and miR-375-3p are consistently enriched in T1DE1 and show an inverse relationship with age. (A–B) Volcano plots from differential expression analyses comparing plasma miRNA profiles between T1D endotypes in two independent cohorts (First Cohort, A; Second Cohort, B). The x-axis reports log2 fold change (log2FC) and the y-axis –log10 adjusted p value (padj); the dashed horizontal line indicates the nominal significance threshold. miR-150-5p and miR-375-3p are highlighted and labeled, showing consistent upregulation in T1DE1 across cohorts. (C–F) Association between visit-1 age (V1_Age) and normalized plasma miRNA counts for miR-150-5p (C, E) and miR-375-3p (D, F) in the First Cohort (C–D) and Second Cohort (E–F). Each dot represents one individual and is color-coded by age strata (<7 years, 7–12 years, ≥13 years). Solid lines indicate the fitted trend. Spearman's rank correlation coefficients (rho) and p values are reported in each panel, showing a significant negative correlation between miRNA abundance and age in both cohorts. Values are fitted on a Log10 scale.

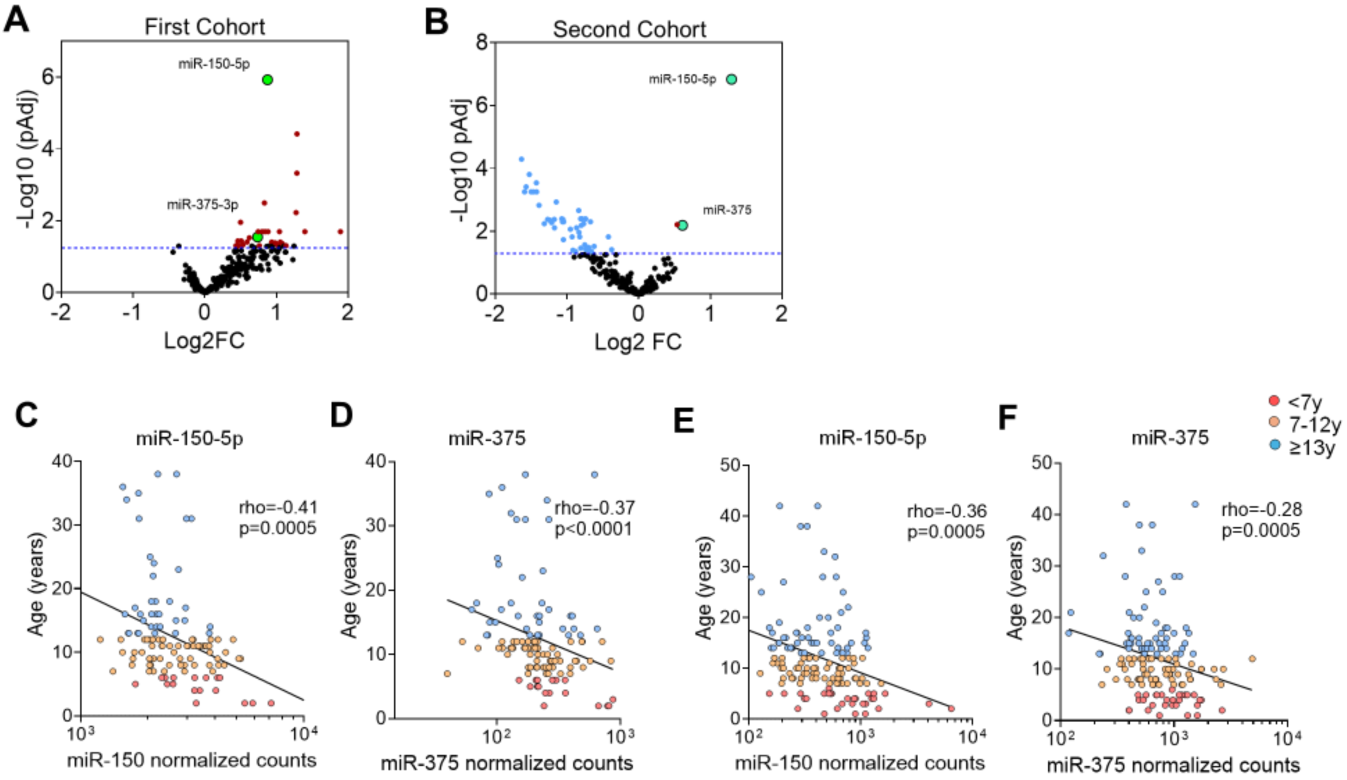

**Supplemental Figure 2.** Circulating miR-150-5p and miR-375-3p levels are not driven by chronological age. Scatterplots show the relationship between age and normalized circulating miRNA counts (log2 scale) for miR-150-5p (A–C) and miR-375-3p (D–F) across three study groups: controls (CTR), children with diabetes (CD), and an independent cohort (CA). Background shading indicates the age strata used in downstream analyses (<7 years, 7–12 years, and ≥13 years). Solid lines depict the fitted trend, and Spearman's rank correlation coefficients (rho) with corresponding p values are reported in each panel. Overall, neither miR-150-5p nor miR-375-3p showed a significant correlation with age within any group, supporting that the observed endotype-associated differences are not simply attributable to age distribution

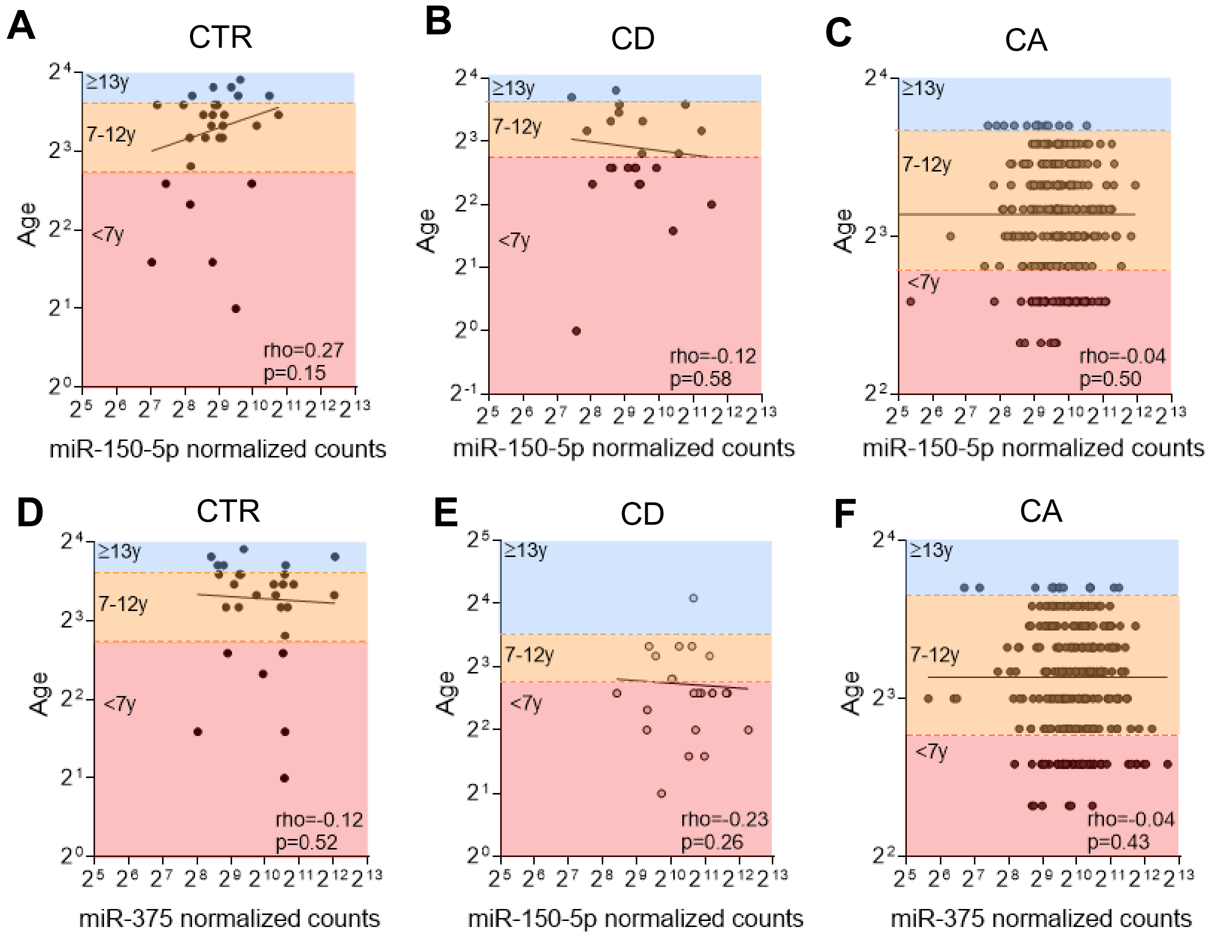

**Supplemental Figure 3.** miR-150-5p is preferentially expressed in lymphoid lineages in a public miRNA-seq atlas. (A) Distribution of miR-150-5p abundance across human tissues and biofluids, reported as reads per million (RPM; log2 scale). Plasma and whole blood are highlighted (red arrows) to contextualize circulating detection relative to tissue expression. (B) Distribution of miR-150-5p expression across primary human cell types (RPM; log2 scale), showing marked enrichment in lymphocyte populations, including $CD8^+$ T lymphocytes, $CD4^+$ T lymphocytes, $CD19^+$ lymphocytes, and B lymphocytes (red arrows; highest-expression groups highlighted in red). Each dot represents an independent sample/dataset; boxplots indicate median and interquartile range, with whiskers reflecting the spread of values.

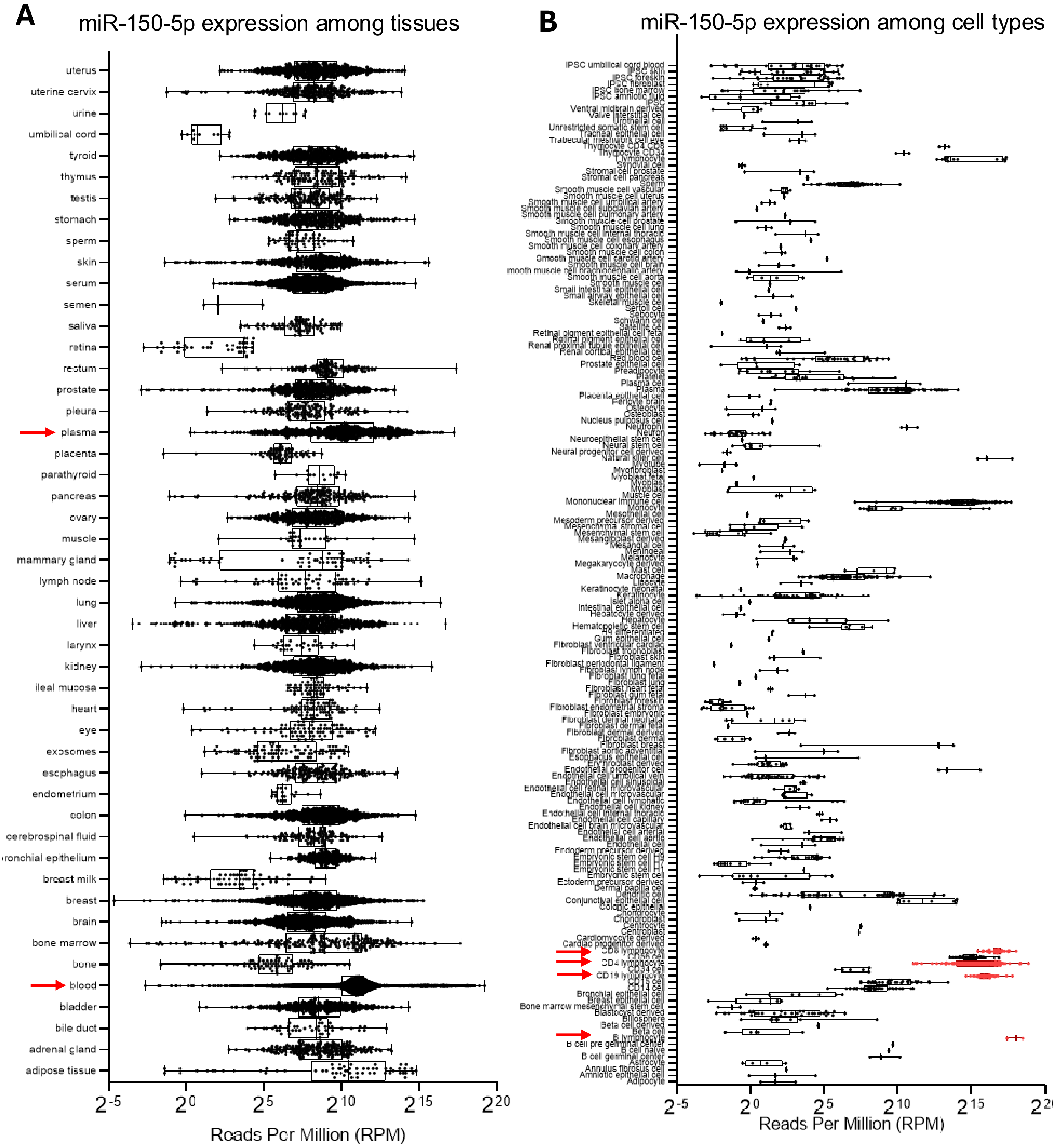

**Supplemental Figure 4.** ROC curves analyses of T1DE1 and T1DE2 individuals of the first and second cohort based on miR-150-5p (**A**) and miR-375-3p (**B**) expression levels (copies/uL). For miR-150-5p a total of n=143 T1D individuals (n=48 T1DE1; n=95 T1DE2) with available ddPCR measurements were included. For miR-375-3p a total of n=135 T1D individuals (n=46 T1DE1; n=89 T1DE2) with available ddPCR measurements were included. For each ROC, AUC and sensitivity and specificity values are reported, alongside to Youden-index ddPCR cutoff and P values.

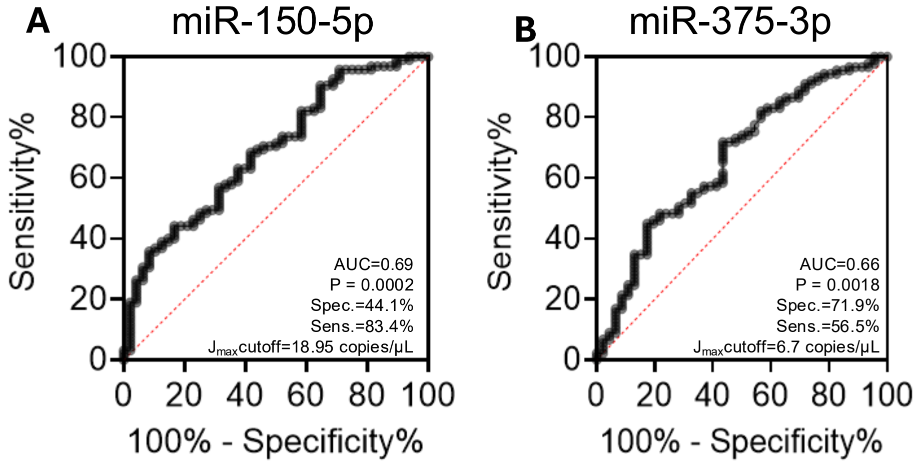
